# Patterns of Childhood Exposomes, White Matter Development and Behaviors Link to Future Cognition and Psychopathology in Youth

**DOI:** 10.64898/2026.11.09.26367658

**Authors:** Danni Wang, Christopher J Hammond, Betty Jo Salmeron, Xiang Xiao, Laura Murray, Hong Gu, Tianye Zhai, Nanyu Kuang, Hui Zheng, Justine Hill, Hanbing Lu, Amy Janes, Thomas J Ross, Yihong Yang

## Abstract

Environmental exposures during childhood strongly impact individuals’ cognitive and socioemotional development, and psychiatric vulnerability. However, childhood exposures rarely occur in isolation. Instead, they form a highly correlated ‘exposome’ whose multidimensional relationships with brain development and behavioral outcomes remain poorly understood. Elucidating the complex interplay between brain, behavior, and environment requires a multivariate approach. This study used regularized multiset canonical correlation analysis on longitudinal data from 8,113 preadolescents in the Adolescent Brain Cognitive Development study to characterize latent multivariate dimensions linking diverse environmental exposures, tract-wisely resolved measures of white matter (WM) development and integrity, and multi-domain behavioral phenotypes and relate them to concurrent and future psychopathology. Two distinct multivariate dimensions were identified exhibiting different exposure, WM development, and behavior patterns. The first linked greater socioeconomic resources to more advanced WM development, particularly in dorsal-association tracts, and superior cognitive performance. The second dimension linked stronger psychosocial support with higher WM integrity, more advanced development of limbic tracts, and lower levels of psychopathology. Longitudinal analyses revealed differential contributions of these dimensions to follow-up cognitive, socioemotional and psychiatric outcomes. Moreover, WM and exposomic variates in these dimensions moderated the effects of childhood adversity on transdiagnostic psychiatric disorder risk, in sex-and context-specific fashions. Together, these findings clarify how childhood environmental exposures and the linked WM developmental differences relate to cognition and psychopathology risk, potentially informing the identification of candidate neural targets for future interventions.

## Introduction

Adolescence is a developmental period typified by neuroplastic brain changes and rapid development of cognitive and socioemotional processing systems, coinciding with increased vulnerability to a wide range of psychiatric disorders (1, 2). During this period, environmental exposures, positive and negative, can collectively and differentially influence developmental trajectories and health outcomes (3). Previous studies using risk-based frameworks have shown that exposure to negative environmental factors such as poverty, community violence, parental substance use have deleterious effects on development and are associated with increased risk for negative health outcomes, including psychiatric disorders (4–6). More recently, resilience-based frameworks have highlighted that exposure to negative environmental factors don’t inevitably lead to adverse outcomes, and that exposures to positive factors such as parental support may buffer against the risk for negative outcomes and promote positive health outcomes (7, 8). Substantial heterogeneity exists in developmental outcomes following childhood exposure to adverse environments, with a large portion of individuals maintaining psychological well-being despite exposure to childhood adversity (7, 9). However, less is known about the neurodevelopmental mechanisms through which complex multidimensional environmental exposures shape both risk and resilience to psychiatric disorders. A major gap in this space is that few studies have examined positive and negative exposures together on health outcomes in the same sample.

White matter (WM) tracts, particularly those supporting cognitive and socioemotional brain systems, undergo protracted and experience-dependent refinement across childhood and adolescence (10–12). These processes position WM development as a key neurobiological substrate through which environmental exposures, positive and negative, become imprinted on brain architecture. Experience-dependent mechanisms, including neuronal firing and stressor-related signaling, regulate oligodendrocyte proliferation, myelin sheath formation, and other cellular mechanisms that influence the development of different WM pathways (10, 13). Importantly, such plasticity enables WM pathways to dynamically adapt to environmental demands, suggesting that WM development may encode both adaptive and maladaptive responses to environmental exposures. Recent advances in normative modeling provide a powerful framework to quantify individual differences in neurodevelopment. Specifically, the brain age gap (BAG), defined as the difference between predicted biological age and chronological age, offers an individualized measure of brain development that captures inter-individual variability beyond chronological age effects. While prior studies have primarily focused on global or whole-brain BAG (14, 15), we recently extended this framework to tract-specific WM BAGs that enable a more precise characterization of neurodevelopmental heterogeneity (16). In this context, tract-specific BAGs quantify deviations from normative developmental trajectories at the level of individual WM pathways, allowing the identification of distinct patterns of accelerated or delayed maturation across distributed brain systems. Such heterogeneity is critical, as different WM tracts subserve distinct cognitive and socioemotional functions and may exhibit differential sensitivity to environmental exposures.

Previous studies have demonstrated that distinct environmental factors can influence WM development in a tract-specific manner. For example, early institutionalization has been shown to alter the microstructure of uncinate fasciculus and corpus callosum (17), and childhood maltreatment has been linked to lower FA in tracts including cingulum and superior longitudinal fasciculus (SLF) (18–20), while higher educational enrichment is associated with higher factional anisotropy (FA) in arcuate fasciculus (AF) and more positive BAG in whole-brain WM during preadolescence (14). However, studies to date have primarily focused on specific environmental exposures, not on exposures in aggregate or considering positive and negative factors together. Additionally, findings from these studies have largely relied on univariate or regional analyses of conventional WM microstructural properties, limiting the ability to capture coordinated patterns of variation across environmental exposures, WM development and behavior. Importantly, environmental exposures rarely occur in isolation. The exposome conceptual framework posits that the totality of environmental exposures (chemical, physical, biological, and social) that an individual experiences across their lifespan (termed the ‘exposome’) interacts with that individual’s genetic architecture to influence biology, functioning, and disease susceptibility (21, 22). Within this framework, socioeconomic, family, school, neighborhood and psychosocial exposures frequently co-occur and are inter-dependent/co-dependent. Thus, neurodevelopmental outcomes are shaped not by single exposures but by multidimensional environmental exposure profiles. Multivariate approaches provide an opportunity to disentangle the latent dimensions capturing different patterns of environmental exposures, brain development, and behavioral phenotypes. To fill this research gap, as the first goal of this study, we conducted a multivariate analysis to identify latent profiles linking childhood environmental exposures, WM development, and cognition and transdiagnostic psychopathology in preadolescence.

Beyond characterizing the concurrent associations, the identified environmental and WM dimensions may reflect broader forms of individuals’ resources and capabilities (23), encompassing cognitive, socioemotional, and resilience-related attributes that contribute to future behavioral outcomes and psychiatric risks. Such predictive dimensions may provide early indicators of risk and resilience before the onset of psychiatric disorders that can be used to guide personalized prevention and early intervention strategies improving population health. However, it remains largely unknown how latent dimensions jointly linking environmental exposures, WM development, and behavioral phenotypes in preadolescence relate to subsequent developmental trajectories and psychopathology risk. The second goal of our study, therefore, is to investigate whether derived neurodevelopmental and environmental signatures contribute risk or resilience to prospectively characterized psychiatric disorders and cognitive and socioemotional outcomes.

In the present study, we introduce a three-way multivariate analytical framework and comprehensively leveraged a wide variety of tract-specific developmental, environmental and behavioral measures in 8,113 participants in Adolescent Brain Cognitive Development (ABCD) Study (24, 25). We applied a regularized multi-set canonical correlation analysis (rmCCA) approach to characterize the covaried dimensions between environment, WM development (16), and behavior based on the baseline data. By incorporating a broadly defined exposome including positive and negative exposures, we were able to test both risk-and resilience-based frameworks, evaluating the implications of these multivariate dimensions in: (1) explaining follow-up cognitive/socioemotional outcomes and transdiagnostic status of psychiatric disorders; and (2) moderating the effects of recent adverse childhood experiences (ACE) on transdiagnostic status of psychiatric disorders.

## Results

### Covarying multidomain dimensions linking white matter development, behavior and environment in preadolescents

To comprehensively characterize neurodevelopment, behavior, and childhood environmental exposures in baseline ABCD participants, we analyzed three complementary data sets: 60 tract-specific WM measures (16), 51 multidomain behavioral measures spanning cognition and psychopathology, and 312 exposome measures capturing family, school/peers, neighborhood, lifestyle, prenatal and substance-related exposures (Figure 1, see Methods for more details). The WM measures were derived from diffusion MRI data harmonized across imaging sites to minimize scanner-related nonbiological variability. WM measures included 30 tract-wise fractional anisotropy (FA) measures characterizing WM microstructure and 30 corresponding FA-derived brain age gaps (BAGs). These tract-specific BAGs were calculated using the tract-specific brain age models developed in our previous study (16) and indexed individual differences in tract-specific measures relative to age-based expectations. We included both FA and BAG because our previous work demonstrated their complementary and differential sensitivity to individual differences relevant to cognitive development and psychopathology (16). After quality control, 8,113 participants (3,921 girls; age range 8.92-11.08 years) with complete WM, behavioral, and environmental data were included in subsequent analyses.

**Figure 1.**
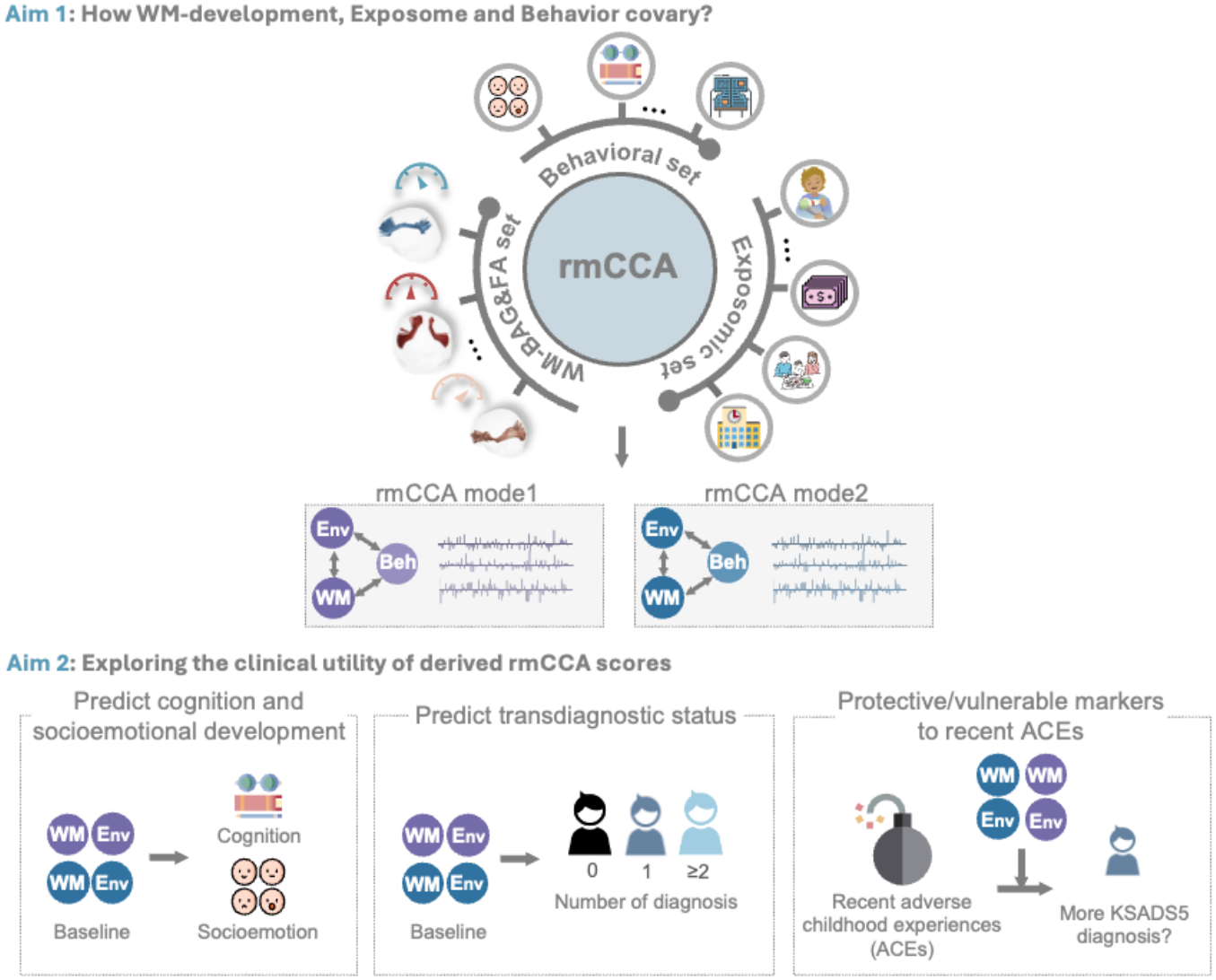
Overview of study design and analytic framework. We applied regularized multi-set canonical correlation analysis (rmCCA) to identify latent dimensions that jointly capture variance across three domains: tract-specific white matter (WM) features, multidimensional environmental exposures, and behavioral assessments. Next, we examined the clinical relevance of the derived WM and environmental latent profiles in predicting three key outcomes: 1) future cognitive and socioemotional development, 2) follow-up transdiagnostic psychiatric risks assessed by the cumulative number of KSADS-5 diagnoses, and 3) moderation effects on the relationship between recent adverse childhood experiences (ACEs) and longitudinal changes in psychiatric diagnoses. Abbreviations: KSADS-5, Kiddie Schedule for Affective Disorders and Schizophrenia for DSM-5. Icons were made from https://www.svgrepo.com/.

To examine how WM development, behavior (Beh) and environment (Env) covary during preadolescence, we applied regularized multiset canonical correlation analysis (rmCCA) to a large population-based cohort (Figure 1). This multivariate approach identified latent dimensions (i.e., modes) that capture maximal covariance across three domains. After 1,000 permutation tests, the significance of each mode was assessed by comparing the observed average inner variance and pairwise covariance with their corresponding null distributions. Z-scores were calculated by dividing the observed values by the standard deviation of the permuted estimates. Two modes were found to be significant, with their average inner variance and pairwise covariances exceeding the null distributions (1^st^ rmCCA variate: *z_all_* = 60.35, *p_all_* < 0.001, *z*_W*M*–*Be*ℎ_ = 29.43, *p*_W*M*–*Be*ℎ_ < 0.001, *z*_W*M*–*Env*_ = 24.74, *p*_W*M*–*Env*_ < 0.001, *z_Env_*_–*Be*ℎ_ = 100.53, *p_Env_*_–*Be*ℎ_ < 0.001 ; 2^nd^ rmCCA variate: *z_all_* = 25.13, *p_all_* < 0.001, *z*_W*M*–*Be*ℎ_ = 15.75, *p*_W*M*–*Be*ℎ_ < 0.001, *z*_W*M*–*Env*_ = 11.24, *p*_W*M*–*Env*_ < 0.001, *z_Env_*_–*Be*ℎ_ = 33.63, *p_Env_*_–*Be*ℎ_ < 0.001; Supplementary Figure S1B-C). The two modes showed distinct patterns of brain-behavior-environment covariation, each linking tract-specific deviations in WM development/integrity to differential behavioral and exposome profiles (Figure 2).

**Figure 2.**
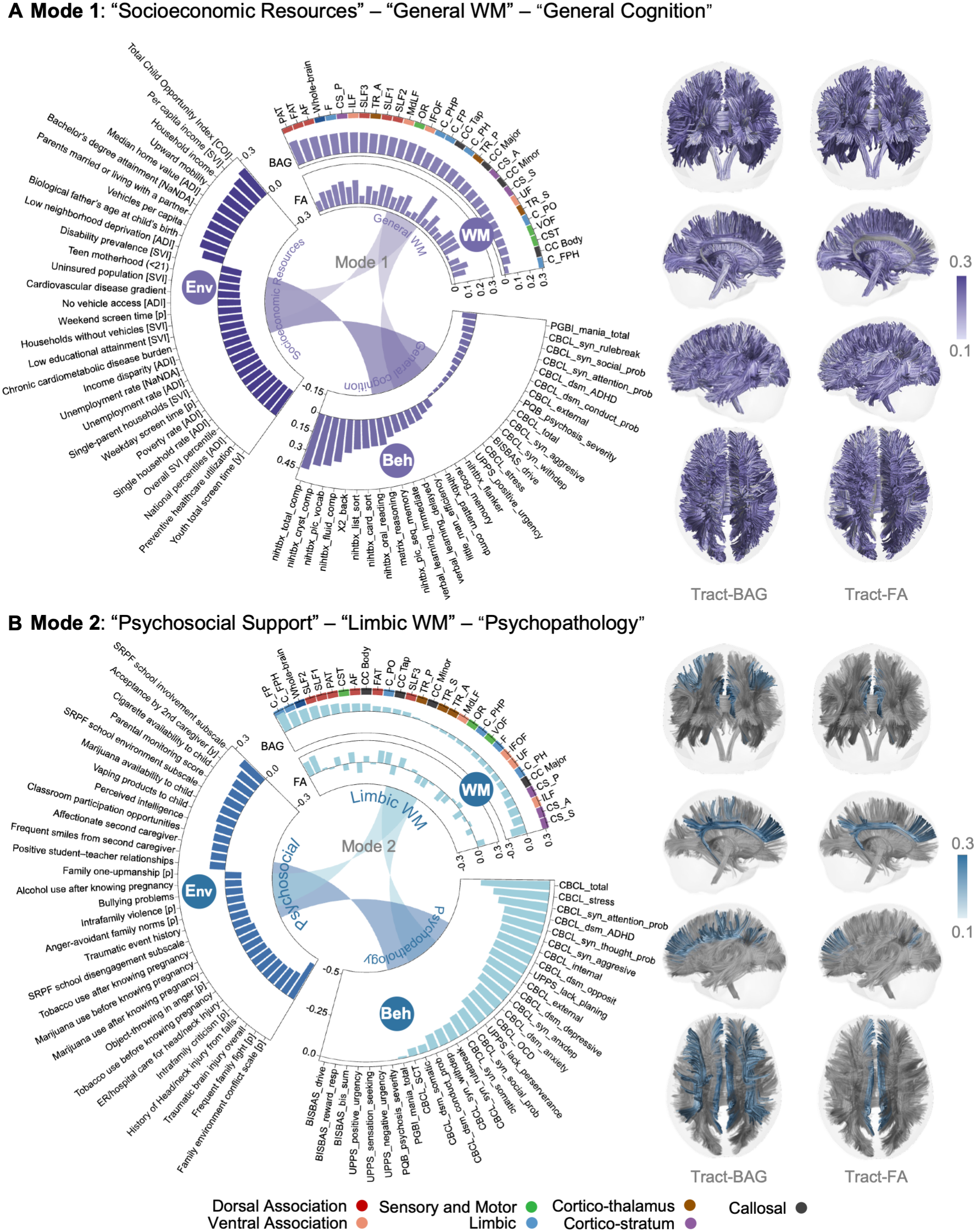
Two covarying multivariate dimensions (modes) of white matter (WM) development, environmental exposures and multi-domain behaviors. Chord plots show the loadings of WM developmental (WM), Environmental (Env), and Behavioral (Beh) data sets for the two significant rmCCA modes. Bar lengths indicate the feature contribution (loadings) of the 30 variables with the largest absolute loadings, while the connecting ribbons represent the pairwise covariation between domains. WM tracts are color-coded by tract system. Brain renderings display the spatial distribution of tract-specific BAG (left) or FA (right) loadings on each rmCCA mode, mapped onto whole-brain WM streamlines. Only WM tracts with bootstrap-significant loadings are displayed. Streamline colors correspond to the loading strength. Abbreviations: BAG, brain age gap; FA, factional anisotropy; ADI, Area Deprivation Index; SVI, Social Vulnerability Index; CRPBI, Children’s Report of Parental Behavioral Inventory; CG2, secondary caregiver; TBI, traumatic brain injury; CBCL, Child Behavioral Checklist; PGBI, Parent General Behavior Inventory; UPPS, Urgency, Perseverance, Premeditation and Sensation-seeking; OCD, obsessive-compulsive disorder; ADHD, attention-deficit/hyperactivity disorder; nihtbx, NIH toolbox. Full variable names of WM tracts are provided in Supplementary Table S1 (document file). Detailed descriptions of the exposomic variables are provided in Supplementary Data S1 (spreadsheet file).

The 1^st^ mode was characterized by increased BAG suggesting more advanced development of WM tracts, particularly in the dorsal association system (e.g. frontal aslant tract [FAT], parietal aslant tract [PAT] and arcuate fasciculus [AF], Figure 2A) and higher FAs in cingulum parietal hippocampal (C_PH) tract after 5,000 bootstrap sampling tests. More positive BAGs and higher FA were associated with better generalized cognitive performance across tasks and domains, lower externalizing problems, higher socioeconomic resources, and lower digital device engagement. The positive canonical loadings for the behavioral domain were mainly characterized by measures of generalized cognition including total and crystalized cognition composite scores. The greatest negative loadings were from the psychopathological measures including child behavioral checklist (CBCL) rule breaking, attention problems, social problems, and parent general behavior inventory (PGBI) total score of mania. Significant environmental contributors included measures indexing higher neighborhood-level child opportunity index (COI), higher household income, less screen time, and lower percentage of areal population below the poverty level (see Supplementary Data 3 and Figure S2-6 for more details). Hereafter, we use the terms General WM Variate and Socioeconomic Resources Variate to describe the WM and environmental canonical variates in the 1^st^ mode, respectively, named based on their strongest loading features.

For the 2^nd^ mode, more positive BAGs of limbic tracts, such as cingulum frontal parietal (C_FP) and cingulum frontal parahipocampal (C_FPH), were the most heavily loaded WM developmental features. These tracts also demonstrated the highest loadings within the WM FA feature set. As shown in Figure 2B, higher brain variate scores in the 2^nd^ mode were linked to lower CBCL total, stress and attention problems and ADHD scores. The environmental measures with the strongest loadings were characterized by school or family environmental measures indexing lower levels of family conflict and fight and prenatal marijuana and tobacco exposure, higher parental monitoring score, greater school involvement, and more positive school environment. See Supplementary Data 3 and Figure S2-6 for more details. Based on these loadings, the WM canonical variate in the 2^nd^ mode is referred to as Limbic WM Variate, while the environmental canonical variate in the 2^nd^ mode is referred to as Psychosocial Support Variate.

### White matter developmental and environmental canonical variates predicted cognitive and socioemotional development one to six years later

Next, we examined whether these baseline WM developmental and environmental canonical variates contribute to cognitive and social abilities at follow-up visits. To ensure independence from the original rmCCA results, we selected follow-up measures that were not included in the previous analysis. The cognitive development measures included math ability measured by Stanford Mental Arithmetic Response Time Evaluation (SMARTE) at 3- and 5-year follow-ups, Emotional Stroop Task Accuracy (STRP Acc) at 1-, 3-, and 5-year follow-ups, and school grades at 2-and 3-year follow-ups. Socioemotional development was evaluated using total scores from the Barkley Deficits in Executive Functioning Scale (BDEFS) at 3-and 6-year-follow-ups, and the Difficulties in Emotion Regulation (DER) measure across five approximately annual follow-up waves, corresponding to approximately 1-5 years after baseline. Associations between each baseline canonical variate (CV) score and each longitudinal outcome were assessed using linear mixed-effects (LME) models, with age and sex as fixed-effect covariates and random intercepts for study site and family.

As shown in Figure 3A, the Socioeconomic Resources Variate scores were the strongest and most significant predictors for all future cognitive measures (e.g. 5-year-follow-up SMARTE: t=27.895, *β* = 0.372, 95% *CI* = [0.346, 0.398], *p_FDR_* < 0.001). In contrast, the Psychosocial Support Variate scores showed statistically significant but much weaker associations with school grades across follow-ups. On the WM side, higher General WM Variate scores were significantly associated with better performance of all cognitive measures at all time points. Multiple comparisons were corrected using the FDR procedure. See detailed statistics in Supplementary Table S2.

**Figure 3.**
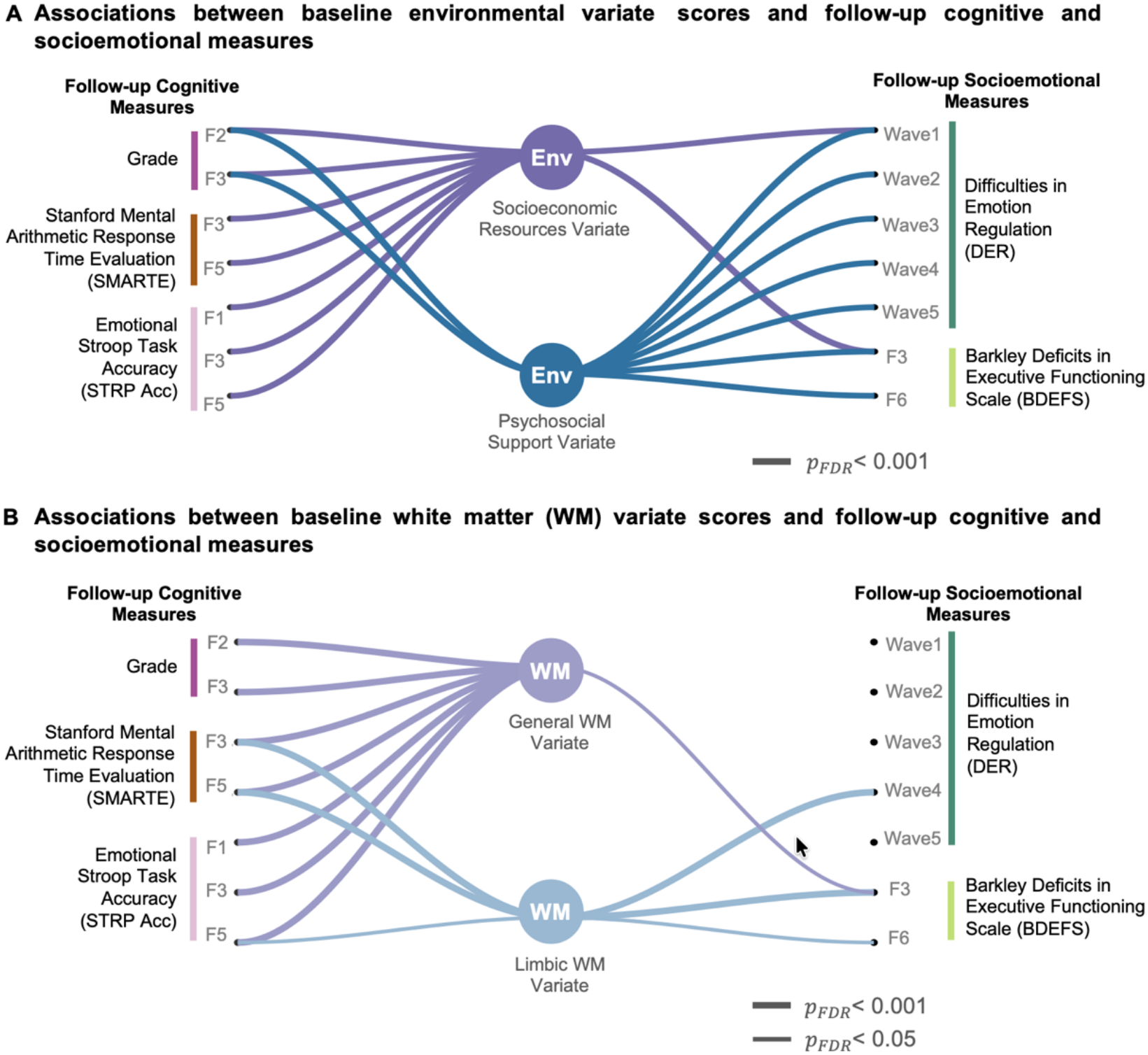
Associations of baseline environmental (Env) and white matter (WM) developmental variate scores with follow-up cognitive and socioemotional outcomes. (A) Associations between baseline environmental variate scores and follow-up cognitive (left panel) and socioemotional (right panel) outcomes. (B) Associations between baseline WM variate scores and the same follow-up outcomes. Only associations that survived false discovery rate (FDR) correction (*p_FDR_* < 0.05) are shown. Thick lines indicate (*p_FDR_* < 0.001), and thin lines indicate (*p_FDR_* < 0.05). Color coding reflects the corresponding latent modes (purple vs blue). Abbreviations: SMARTE, Stanford Mental Arithmetic Response Time Evaluation; DER, Difficulties in Emotion Regulation Scale; BDEFS, Barkley Deficits in Executive Functioning Scale; STRP, overall performance of Emotional Stroop task; F, Follow-up. “Follow-up” denotes scheduled years since the ABCD baseline assessment, whereas “Wave” denotes the assessment rounds of the ABCD Social Development substudy, which follows a separate assessment schedule.

For socioemotional development, Figure 3B illustrates distinct longitudinal associations between the two environmental dimensions and follow-up BDEFS and DER measures. The Socioeconomic Resources Variate score only demonstrated statistical significance at the 1^st^ follow-up visits (DER at the 1^st^ visit: *t*=-4.713; *β* = −0.114, 95% *CI* = [−0.161, −0.066]; *p_FDR_* < 0.001). In contrast, the Psychosocial Support Variate score consistently showed the most significance in DER and BDEFS scores across nearly all follow-up visits. On the WM side, the Limbic WM Variate explained more variance, compared to the General WM Variate, for follow-up socioemotional outcomes (e.g. BDEFS at the 3-y-follow-up: *t*=-3.564; *β* = −0.042; 95% CI: [-0.065, -0.019]; *p_FDR_* < 0.001; DER at the 4^th^ visit: *t*=-3.963; *β* = −0.105; 95% CI: [-0.156, -0.052]; *p_FDR_* < 0.001). See more detailed statistics in Supplementary Table S3.

### White matter developmental and environmental canonical variates at baseline associated with concurrent and prospective cumulative number of psychiatric diagnoses

We next examined whether environmental and WM developmental canonical variates were associated with the cumulative number of psychiatric diagnoses both concurrently and at 2-year-follow-up. We used the baseline and 2-year K-SADS-5 assessments so that the interval for measuring changes in psychiatric diagnoses matched the interval used to quantify changes in recent adverse childhood experience (ACE) scores in the following moderation analysis. LME models compared baseline canonical variate scores across K-SADS-5 diagnostic groups, with age and sex as fixed-effect covariates and random intercepts for study site and family ID. As shown in Figure 4A, higher canonical variate scores of both environmental modes were significantly associated with fewer cumulative KSADS-5 diagnoses concurrently and at the 2-year-follow-up. Of note, the variance explained by Socioeconomic Resources Variate decreased over time (Baseline: *F* = 37.567, *p_FDR_* < 0.001; 2-y-follow-up: *F* = 11.085, *p_FDR_* < 0.001), whereas the Psychosocial Support Variate was stable (Baseline: *F* = 144.405, *p_FDR_* < 0.001; 2-y-follow-up: *F* = 103.766, *p_FDR_* < 0.001). For the WM variates, the General WM Variate score was significantly and negatively associated with the cumulative number of psychiatric diagnoses at baseline and 2-y-follow-up visits (Baseline: *F* = 5.631, *p_FDR_* = 0.0048; 2-y-follow-up: *F* = 6.950, *p_FDR_* = 0.0013). In post-hoc group comparisons, the General WM Variate score was significantly lower in individuals with 2 or more diagnoses compared to those without diagnoses concurrently and at 2-year-follow-up.

**Figure 4.**
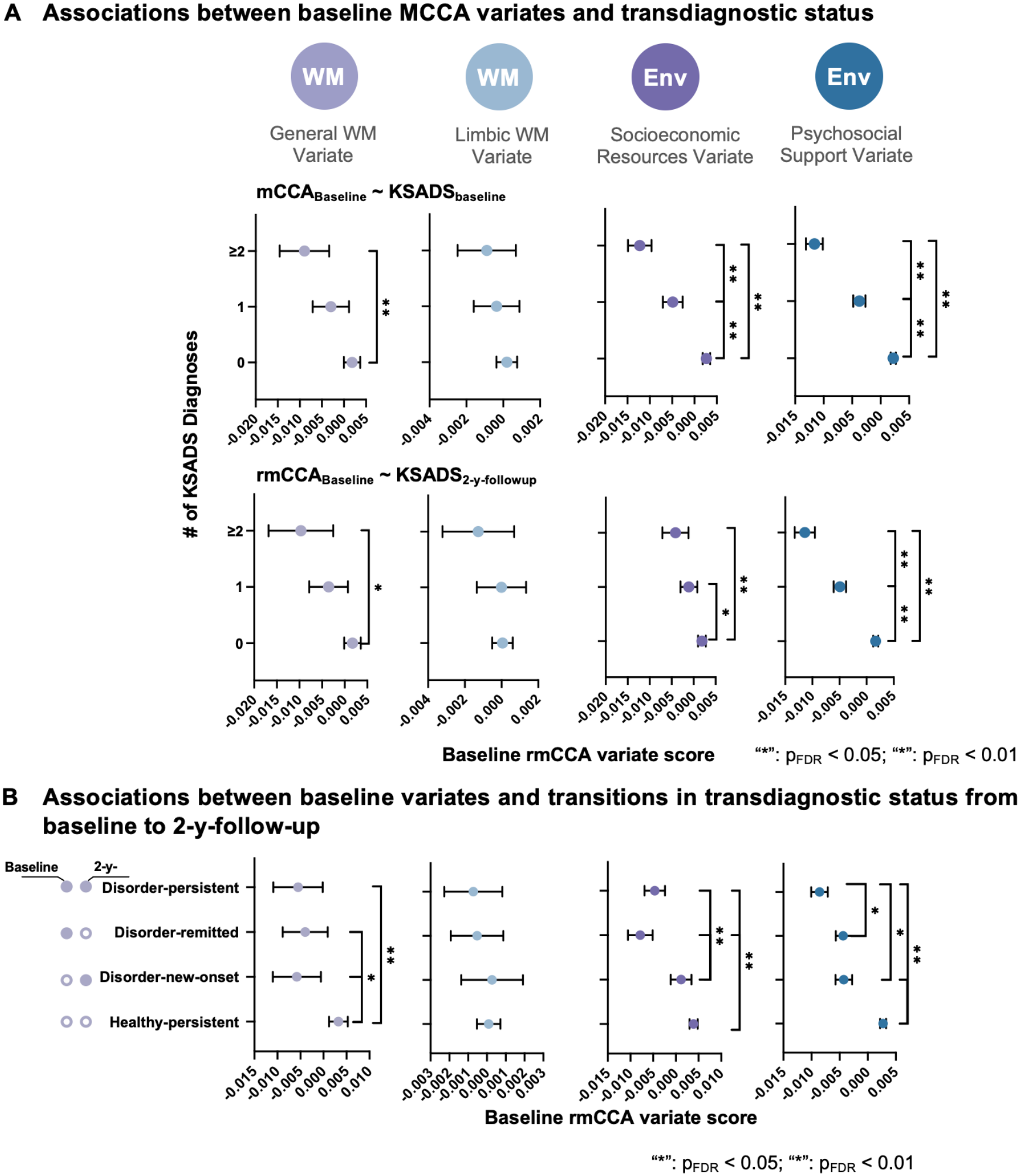
Associations between baseline white matter (WM) and environmental variate scores and longitudinal transdiagnostic psychiatric risks. (A) Group-level comparison of the baseline WM and environmental variate scores and the cumulative numbers of diagnosis at baseline (top panel) and 2-year-follow-up (bottom panel) as measured by the KSADS-5. (B) Association between baseline variate scores and transitions in transdiagnostic status from baseline to 2-year-follow-up. The participants were categorized into four subgroups: Healthy-persistent, Disorder-remitted, Disorder-new-onset and Disorder-persistent group. Error bars represent 95% confidence intervals. “*” *p_FDR_* < 0.05; “**” *p_FDR_* < 0.01. Abbreviations: KSADS-5, Kiddie Schedule for Affective Disorders and Schizophrenia for DSM-5.

We also investigated associations of the scores of the four canonical variates at baseline with the psychiatric diagnostic trajectory (e.g., Healthy-persistent, no psychiatric diagnosis for both visits; Disorder-remitted, ≥1 psychiatric diagnosis at baseline and no diagnosis at 2-year follow-up; Disorder-new-onset, no psychiatric diagnosis at baseline and ≥1 diagnosis at 2-year follow-up; and Disorder-persistent, ≥1 psychiatric diagnosis for both visits) from baseline to 2-year follow-up. As shown in Figure 4B, the two environmental variate scores and the General WM Variate scores showed significant associations with diagnostic trajectories. Post-hoc subgroup-wise comparisons showed that the scores of all three Variates were significantly lower in the Disorder-persistent subgroup compared to the Healthy-persistent subgroup. Furthermore, the Socioeconomic Resources Variate scores were significantly lower in the subgroups with baseline diagnoses compared to the subgroups without baseline diagnoses, while the Psychosocial Support Variate showed lower canonical variate scores in subgroups with follow-up diagnoses compared to the ones without follow-up diagnoses, including healthy-persistent versus disorder-new-onset, patient-remitted versus patient-persistent subgroups.

### Environmental and WM developmental variates buffer against the impacts of recent adverse childhood experiences (ACEs) on developing transdiagnostic psychiatric disorders in adolescents

Finally, we examined whether baseline environmental and WM developmental variates have protective effects against the impacts of recent ACEs on the development of psychiatric disorders (Figure 5A). As shown in Figure 5B, higher recent ACEs during the two years from baseline to 2-year-follow-up were significantly associated with larger changes in cumulative number of total KSADS-5 diagnoses (*rho* = 0.146, *p_FDR_* < 0.001) across the two years. Moderation analyses revealed that General WM Variate scores (*t* = -2.620, *p_FDR_* = 0.012), as well as Socioeconomic Resources (*t* = -4.168, *p_FDR_* < 0.001) and Psychosocial Support Variates (*t* = -3.208, *p_FDR_* = 0.003), significantly attenuated the association between recent ACEs exposure and transdiagnostic risks (Figure 5C). We further examined whether these moderation effects differed by sex. As shown in Figure 5D, the moderation effect of General WM Variate score on the association between recent ACEs and changes in KSADS-5 diagnoses differed significantly by sex (*t* = 2.297, *p* = 0.022), with significant moderation effect in girls (*t* = -3.185, p = 0.001), but not in boys (*t* = -0.393, *p* = 0.694). However, the other canonical variates did not show significant sex difference in the moderation analysis. See other moderation results in Supplementary Table S4.

**Figure 5.**
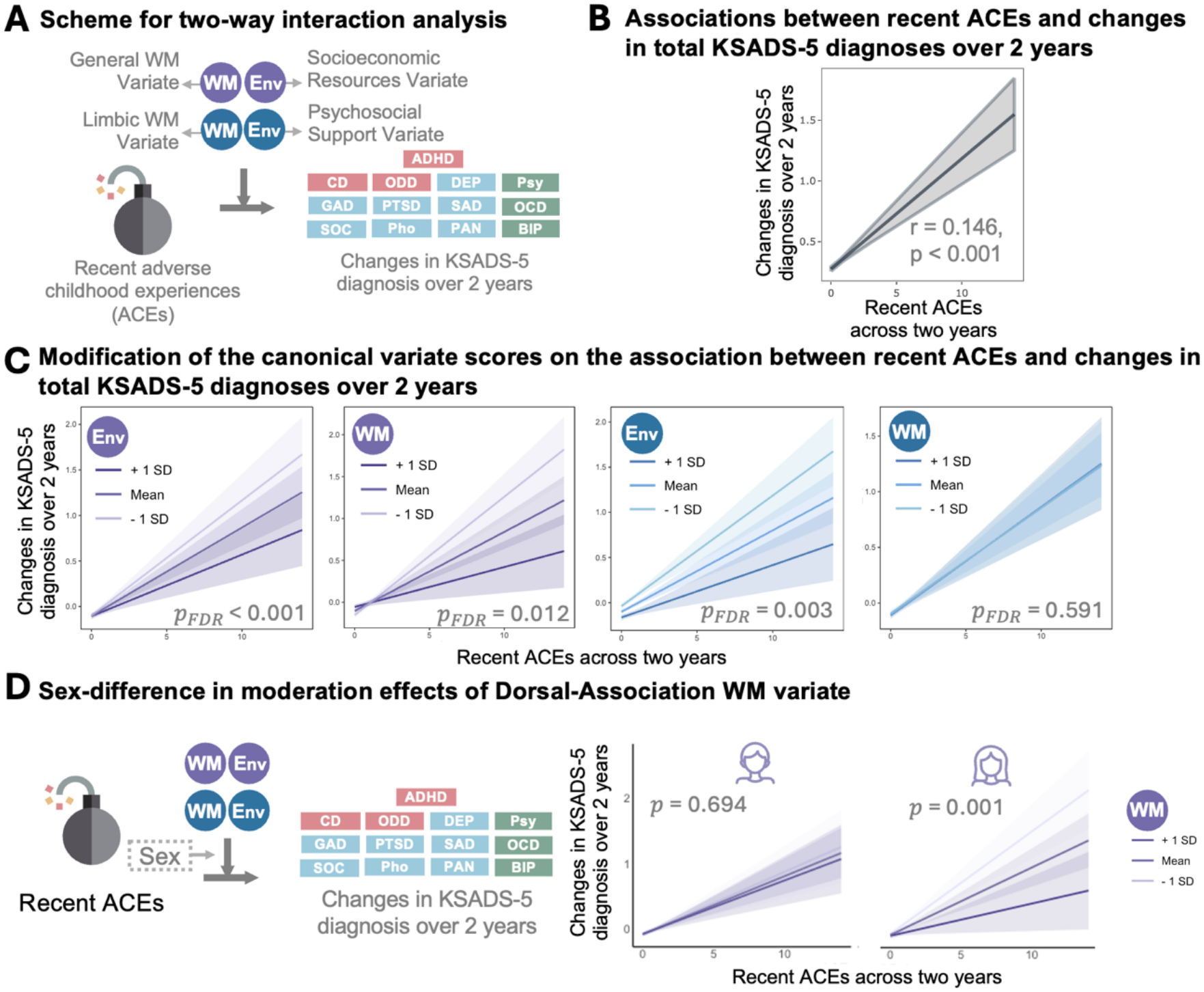
Environmental (Env) and white matter (WM) canonical variate scores modulate the impact of recent adverse childhood experiences (ACEs) on psychiatric diagnoses. (A) Schematic overview of the two-way interaction analysis. (B) Association between recent ACEs and changes in total KSADS-5 diagnoses over the two years. (C) Moderation effects of environmental and WM variate scores on the association between recent ACEs and changes in KSADS-5 diagnoses. (D) Sex-specific moderation effects of variates on the association between recent ACEs and changes in total KSADS-5 diagnoses. The left panel illustrates the three-way interaction model. The right panels show that General WM variate significantly moderated the association between recent ACEs and changes in KSADS-5 diagnoses over two years in girls but not boys. Lines represent simple slopes at +1 SD, mean, and -1 SD of the moderator scores; shaded regions indicate 95% confidence intervals. Abbreviations: KSADS-5, Kiddie Schedule for Affective Disorders and Schizophrenia for DSM-5; rmCCA, regularized multiset canonical correlation analysis.

To comprehensively characterize context-dependent resilience, we conducted a four-way interaction analysis that integrated recent ACEs, WM variates, and environmental contexts (Supplementary Figure 7A). As illustrated in Supplementary Figure 12B, significant higher-order interactions were observed, indicating that the buffering effects of WM variates depended on the surrounding environmental conditions. Specifically, higher General WM Variate scores attenuated the relationship between recent ACEs and increased KSADS diagnoses (t = 3.169, *p_FDR_* = 0.003), particularly in contexts characterized by low socioeconomic resources and limited psychosocial support (Supplementary Figure 7B, left). In contrast, the Limbic WM Variate demonstrated a context-dependent pattern (t = 2.497, *p_FDR_* = 0.013), in which higher scores buffered the adverse effects of recent ACEs in environments with sufficient socioeconomic resources but limited psychosocial support, yet were associated with a stronger recent-ACEs-diagnosis relationship under conditions of more environmental disadvantage on both dimensions (Supplementary Figure 7B, right).

## Discussion

Understanding how brain development, exposome, and behavior interact remains a central challenge in developmental neuroscience. A key difficulty arises from the high dimensionality and strong collinearity within each domain, making it challenging to disentangle how distinct environmental exposures differentially shape neurodevelopmental markers and downstream behavioral outcomes. In this study, we developed a multi-view framework to identify dissociable dimensions linking distinct environmental exposures to different tract-wise developmental and behavioral domains. Together, our findings suggest that environmental influences on WM development are organized along differential system-wise pathways, which in turn confer differential risk and resilience to later transdiagnostic psychopathology.

### Three-way multivariate framework for interrelated environment-WM-behavior relationships

Prior work has primarily focused on examining the relationships between environmental exposures and WM properties in isolation (1, 26), typically focusing on individual exposures and raw tract-level metrics. In contrast, our study extends the tract-wise BAG framework by integrating comprehensive environmental profiles with WM developmental deviations and behavioral outcomes within a unified analytical framework. This three-way framework enables the identification of latent dimensions that jointly capture covariance across environment, neurodevelopmental, and behavioral outcomes.

We identified two significant, mathematically orthogonal dimensions linking environmental exposures, WM development, and behavior. The first dimension was primarily characterized by socioeconomic resources, whereas the second dimension was dominated by psychosocial support. Although these environmental domains largely co-occur in early life (21, 27, 28), our multivariate framework disentangles their distinct contributions by leveraging their shared and unique covariance structures. This allows us to comprehensively characterize tract-level deviations associated with differential exposures that may be obscured in single-exposure or univariate analyses. By linking these different environmental profiles to distinct tract-specific measures and downstream behavioral outcomes, our framework provides a more precise characterization of how specific early-life exposures link to differential neurodevelopmental trajectories. Such differentiation is critical for informing targeted intervention strategies, as it highlights that different environments may shape the developing brain in distinct ways. Moreover, incorporating tract-wise WM BAGs alongside conventional diffusion metrics such as FA offers complementary biological insight. While FA captures microstructural variability along tracts and reflects individual-specific “fingerprints” (29, 30), it does not directly index deviation from normative developmental trajectories. In contrast, tract-wise BAG provides an interpretable measure of developmental deviation, quantifying whether a specific tract exhibits accelerated or delayed maturation relative to age expectations. Together, the integration of multidimensional environmental exposures with tract-specific developmental deviation metrics enables an improved mechanistic understanding of how diverse early-life experiences become embedded in the developing brain and shape behavioral outcomes.

### Two dimensions linking WM development to distinct environmental exposures and behaviors

A central finding of this study is the identification of two orthogonal multivariate modes linking exposome, WM development, and behavior. The first mode is dominated by measures indexing neighborhood and family socioeconomic resources (e.g., Child Opportunity Index, household income), along with lifestyle patterns characterized by less digital engagement and more participation in physical and extracurricular activities. This pattern is consistent with prior work showing that higher socioeconomic status is associated with less screen time and more engagement in physical and extracurricular activities (31, 32). Consistent with recent findings using GM morphological and resting-state functional connectivity neuromarkers (33), socioeconomic exposome measures showed the strongest brain-environment associations; our results extended the importance of socioeconomic exposome to WM development. For the WM dimension, this mode is characterized by more positive tract-wise BAGs and higher FA, suggesting more advanced or efficient WM organization. This aligns with extensive evidence linking socioeconomic advantage to more integral WM microstructure. For example, focal associations between socioeconomic factors and specific tracts demonstrated that higher socioeconomic factors (e.g., household income, parental education) were associated with higher FA in a range of WM tracts like the parahippocampal cingulum and superior cortico-striatum tracts in children and adults (34). More recent studies using the ABCD dataset also showed that higher annual household income was related to higher FAs in SLF and striato-frontal tracts (35). Meanwhile, Roy et al. showed that educational environment is uniquely linked to higher FAs in AF, and more accelerated development of whole-brain WM tracts through brain age modeling (14). Complementary evidence from ABCD study indicated that the variance of longitudinal changes in tract FA differs across household income and caregiver education groups (36). Given that WM development during childhood and adolescence is marked by rapid myelination and axonal refinement, our findings suggest that enriched environments may support more optimal maturation of large-scale association systems. Notably, the strongest BAG loadings were observed in dorsal association tracts, which form the structural backbone of higher-order cognitive networks (16). Consistent with prior literature linking socioeconomic resources to cognitive development (27), this mode mainly captures a latent dimension spanning environmental enrichment, WM organization, and cognitive performance.

In contrast, the second mode reflects a psychosocial dimension characterized by lower family conflict, reduced trauma exposure, greater parental warmth, stronger school engagement, and reduced substance exposure. This mode was associated with a broad transdiagnostic psychopathology profile, with the strongest loadings on total psychopathology, perceived stress, attention problems, ADHD symptoms and thought problems. These behavioral measures covaried with more negative BAGs and lower FAs especially in tracts of frontal-parietal cingulum in preadolescents. Cingulum tract system connects the key fronto-limbic regions (e.g. anterior cingulate cortex and medial prefrontal cortex) within hypothalamic-pituitary-adrenal (HPA) axis. Converging evidence from both preclinical and clinical studies indicate that these circuits are particularly vulnerable to chronic stress (37–39). For instance, heightened cortisol reactivity in children has been associated with reduced FA in WM regions adjacent to the anterior cingulate cortex and medial prefrontal cortex compared to their low-reactive peers (40). Mechanistically, prolonged glucocorticoid exposure has been shown to inhibit oligodendrocyte proliferation and reduce myelination in a dose-dependent manner in rodent models, whereas acute cortisol responses may play adaptive roles in neural plasticity and oligodendrocyte function. Given that our measures reflect environmental exposures from prenatal stages (e.g., prenatal substance and alcohol use) through childhood collected at the ABCD baseline visit, the observed associations likely capture long-term stress-related effects on WM development. The convergence of strong loadings across both FA and BAG dimensions further supports the interpretation that this mode reflects sustained neurodevelopmental deviation associated with chronic stress exposure.

Together, these findings delineate two parallel neurodevelopmental pathways, an enrichment-related pathway supporting cognitive development through dorsal association systems, and a stress-related pathway linked to psychopathology through fronto-limbic circuitry. This framework highlights that environmental influences on brain development are system-specific rather than uniform, with distinct exposures preferentially shaping different WM pathways.

### Differential contributions of the two dimensions to the development of future cognitive and social functions, and psychiatric disorders

It is well established that each individual carries a cumulative imprint of environmental exposures, reflecting a developmental fingerprint shaped over time. However, the relationship between environmental experiences and behavioral outcomes is often complex, as exposures tend co-occur and interact across multiple dimensions. In our results, the two identified modes (or dimensions in the multi-dimensional space) showed strikingly distinct longitudinal associations with behavioral and socioemotional development.

Notably, environmental canonical variates explained a greater proportion of variance in both behavioral outcomes and psychiatric risk than WM-derived variates. The comparatively modest predictive power of neuroimaging markers is consistent with prior work (41, 42) and likely reflects the fact that environment influences multiple biological dimensions, including metabolic, functional, and structural domains (43). Future work integrating multimodal imaging approaches (44) (e.g., myelin water imaging, quantitative susceptibility mapping) may provide a more comprehensive characterization of how environmental exposures are biologically embedded in the developing brain. Identification of the specific tracts linked to differential environmental exposure and linked to future outcomes is important to develop early interventions to resist the negative influence of existing environmental exposures.

Importantly, the two environmental variates showed distinct temporal patterns in their associations with transdiagnostic risks of psychiatric disorders. The Socioeconomic Resources Variate exhibited a diminishing effect on cumulative KSADS-5 diagnoses over time, whereas the Psychosocial Support Variate demonstrated more enduring associations to 2-year follow-up. Consistent with prior literature identifying both socioeconomic disadvantage and adverse psychosocial environments as transdiagnostic risk factors (45), our results further highlight the particularly sustained impact of early-life psychosocial adversity on psychiatric risk. Together, our results clarify that distinct environmental dimensions contribute differentially to developmental outcomes, with enrichment-related processes supporting long-term cognitive trajectories and psychosocial adversity exerting lasting effects on socioemotional and psychiatric development.

### Interactive effects of environmental and WM latent dimensions in shaping resilience under acute adversity

Identifying environmental and neurobiological markers of resilience prior to the onset of psychopathology is essential for informing preventive interventions. Distinct from a risk-based framework, a resilience-based framework emphasizes that exposure to adversity does not inevitably result in psychopathology, and instead highlights the importance of protective processes that promote adaptive outcomes despite risk (7). Consistent with this framework, although greater accumulation of adverse childhood experiences over two years was associated with increased KSADS-5 diagnoses at the population level, individuals with distinct early-life environmental and WM profiles exhibited markedly different susceptibility to transdiagnostic psychiatric risk.

At the exposome level, our findings suggest that more socioeconomic resources and psychosocial support may serve as pre-existing protective factors, reinforcing the idea that positive environmental exposures enhance resilience and facilitate stress regulation (46). The Socioeconomic Resources Variate captured both material and extracurricular enrichment factors, which may mainly promote resilience by cognitive-control development and problem-solving ability to cope with stressor (47). In contrast, the Psychosocial Support Variate was characterized by more family support and positive school environments, which may foster resilience through emotion regulation in response to stress. These findings align with contemporary resilience frameworks proposing that resilience emerges from multiple environmental latent profiles rather than from single protective factor (7).

In addition, our results also indicate that resilience-promoting environmental contexts are embedded in WM systems. The General WM Variate, which was linked to socioeconomic resources and showed the strongest BAG loadings in dorsal association tracts, also buffered the adverse effect of recent ACEs on future psychopathology. The WM tracts within the dorsal association system connect cognitive control networks (16), which are critical for flexible behavioral regulation and goal-directed responses to stress (26). Of note, the buffering effect of the General WM Variate was observed specifically in girls. This sex-specific association may reflect differential strategies to cope with stressful situation between girls and boys, as adolescent girls may be more likely to engage problem-solving and social-support-seeking behaviors when facing adversity, whereas boys more frequently adopt avoidant coping strategies (48). Consequently, individual developmental differences in dorsal association WM system, which supports cognitive control and adaptive coping may buffer greater adverse effect of recent ACEs in girls.

Moreover, our four-way interaction analyses provide evidence that resilience to early-life adversity is not a fixed trait but a context-dependent process arising from interactions between distinct environmental exposures and circuits. Within this framework, the dorsal association WM system may represent a domain-general resilience backbone that supports adaptive functioning even under the more chronic stress from both socioeconomic and psychosocial resources. These tracts underlie cognitive control networks that are critical for flexible behavioral regulation and goal-directed responses to stress (49). Converging functional evidence indicates that disruptions in these networks (50), particularly reduced prefrontal engagement during cognitive control tasks, are associated with exposure to early threat, deprivation, and poverty, highlighting their relevance as potential intervention targets. In addition, our prior finding (16) that the General WM Variate, which showed highly spatial consistency with the General WM Variate in this study (Supplementary Figure 9, spatial correlation R = 0.844), aligns with spatial patterns of mitochondrial content raises the possibility that cellular energy metabolism may represent a mechanistic pathway linking environmental adversity to large-scale network integrity. Indeed, early-life deprivation, such as malnutrition, has been shown to impair mitochondrial function, which may contribute to altered neurodevelopment and reduced resilience (51).

In contrast, limbic pathways appear to support a more context-dependent form of resilience (52), exerting protective effects primarily under conditions of sufficient external resources. In our study, the resilience-promoting effect of the Limbic WM Variate on acute adversity was observed among individuals with relatively adequate socioeconomic resources but limited psychosocial support. This pattern suggests that preserved fronto-limbic integrity may buffer the impact of acute stress when baseline material resources are available, consistent with models of emotion regulation involving top-down control of limbic reactivity (46, 49). At the same time, individuals whose limbic tracts show larger vulnerability, under psychosocial stress but adequate resources, may be better to regulate responses to subsequent stressors. In contrast, deprivation of both socioeconomic and psychosocial resources may create a more severe chronic stress context that reduces the capacity of the regulatory systems to cope with subsequent acute stressors (46). Under such conditions, prior work suggests that stress-related neurodevelopment may shift toward maladaptive trajectories (52), including accelerated maturation of limbic systems (53), which has been linked to heightened emotional reactivity and increased risk for psychopathology.

Together, these findings support a multidimensional model of resilience in which environmental resources and tract-wise systems jointly shape individual differences in adaptation following adversity. Dorsal association pathways appear to provide a domain-general resilience backbone that supports adaptive functioning across diverse environmental contexts, whereas limbic pathways confer more conditional protection that depends on the availability of external resources. More broadly, our results suggest that resilience to psychiatric risks emerges from dynamic interactions between environmental and neurodevelopmental systems rather than from any single protective factor.

## Limitations

This study also has several limitations. First, although the ABCD dataset represents one of the largest developmental cohorts integrating exposome, behavioral, and neuroimaging measures, the present analyses are based on baseline data collected in preadolescents aged 8.92-11.08 years. Environmental measures included retrospectively reported prenatal exposures and childhood environments, with limited information on exposure timing, duration, and accumulation. Although we examined associations with subsequent outcomes, this design limits causal inference and identification of sensitive developmental periods (1). Prospective cohorts with exposome assessments beginning earlier in life, such as the HEALthy Brain and Child Development study (54), could help clarify these temporal relationships. Second, the socioeconomic level of the ABCD cohort is skewed toward higher status relative to the general worldwide population (55). Future studies incorporating developmental datasets with a broader and more diverse range of socioeconomic adversity are necessary to evaluate the generalizability and specificity of the identified modes. Third, environment influences WM development through multiple biological processes, including myelination, neuroinflammation, and glial proliferation. We selected FA based on its robust fingerprinting properties across datasets (29) and the complementary sensitivity of FA and FA-derived BAGs to individual differences in cognition and psychopathology (16). However, both measures derive from the same diffusion metric and have limited biological specificity. Integrating myelin-sensitive or neuroinflammatory imaging (44), would provide a more comprehensive characterization of WM development and may enhance the explanatory power of exposome-brain-behavior associations.

## Conclusions

In summary, we developed a multi-view framework to characterize the complex relationships among the exposome, WM development and behavioral phenotypes. This approach provides a comprehensive perspective on how multidimensional environmental exposures become differentially embedded in WM developmental pathways and are linked to distinct cognitive and psychopathologic outcomes. We identified two distinct exposome-WM-behavior dimensions: one linking socioeconomic resources to general WM development and cognition, and the other linking psychosocial support to limbic WM development, socioemotional functioning, and reduced psychopathology. Furthermore, we demonstrated that environmental exposure patterns and their associated WM pathways contribute to individual differences in vulnerability and resilience to subsequent psychopathology risks. Together, these findings suggest that positive and negative environmental exposures which interactively make up the exposome may become biologically embedded within dissociable WM developmental systems that support distinct domains of adolescent functioning during the peri-adolescent period. Collectively, our findings establish a multilevel neurodevelopmental framework linking environmental experiences, WM development, and psychiatric risks, providing early evidence for identification of vulnerability and resilience factors before the onset of psychiatric disorders.

## Materials and Methods

This cohort study used baseline neuroimaging data and longitudinal behavioral and clinical assessments collected from year-1 to year-5 of the ABCD Study. Reporting followed the Strengthening the Reporting of Observational Studies in Epidemiology (STROBE) guidelines for cohort studies. The ABCD Study was approved by both central and site-specific institutional review boards, and written informed consent and assent were obtained by the ABCD Study teams. Detailed procedures have been described previously. No additional institutional review board approval was required for the present study in accordance with the Common Rule.

### Participants and Study Design

This study used data from the ongoing longitudinal Adolescent Brain Cognitive Development (ABCD) Study (24), which recruited children across 21 US research sites. We included baseline data (release 6.1) from 11,875 children aged 9-11 years, including the imaging data, behavioral and environmental assessments to establish covarying triple-domain relationships. Longitudinal follow-up data were used to examine psychiatric diagnoses as well as cognitive and social development over a period of up to six years.

### Measures

#### White matter developmental and behavioral measures

In our previous study (16), fractional anisotropy (FA) measures along the tract were first harmonized across imaging sites using ComBat (https://github.com/Jfortin1/ComBatHarmonization/tree/master/Matlab) method (56) to reduce nonbiological imaging variability arising from differences in scanners, while preserving biological variation associated with age, sex, and baseline KSADS diagnoses. These harmonized FA measures were subsequently used to develop normative brain age models for 30 tract-specific WM tracts, and to derive tract-specific deviations in WM development via brain age gaps (BAGs) for each participant. In this study, these 30 tract-specific BAG measures were included, indexing WM developmental deviation, alongside 30 corresponding tract-wise FA values representing WM microstructural integrity. In addition, we used 51 behavioral variables (based on our previous studies (16, 57), covering multidomain cognition and psychopathology, measured at baseline as the behavioral data set. Full names and abbreviation of all WM tracts are provided in Supplementary Table 1.

#### Environmental and lifestyle measures

To comprehensively characterize each participant’s environment and lifestyle, we extracted variables from all relevant ABCD parent- and child-reported questionnaires and census-linked external data (25) collected at the baseline visit. Our data preprocessing followed the procedure in previous studies (58, 59). An initial set of 1,154 baseline measures covering domains such as family, school, neighborhood, lifestyle and interpersonal experiences were initially included in the analysis. Variables were excluded if they had more than 10% missing values, less than 1% nonzero values, or no variance. To handle multicollinearity between measures, we aggregated or excluded the highly correlated measures (*R*s > 0.75) within the same questionnaire (59). For example, items of “*During the pregnancy with this child, did you/biological mother have pre-eclampsia, eclampsia, or toxemia?*” and “*During the pregnancy with this child, did you/biological mother have pregnancy-related high blood pressure?*” were aggregated due to highly inter-item correlation coefficient. After data filtering and aggregation, a total of 312 exposome measures were retained for further analysis. Missing values were imputed via the *missForest* library in R (4.6.0). Detailed descriptions of all exposomic measures are provided in Supplementary Data 1.

#### Follow-up assessments for cognitive and social development

To examine how early environmental exposome and associated WM development contribute to follow-up cognitive and social development, we further included the follow-up measures that were independent from those used in the regularized multi-set canonical correlation (rmCCA) analysis. Cognitive development was assessed using math ability (measured by Stanford Mental Arithmetic Response Time Evaluation [SMARTE]) at the 3-year-and 5-year-follow-ups; the overall performance on the Emotional Stroop task (60) at the 1-year-, 3-year-and 5-year-follow-ups; and self-reported school grades at the 2- year-and 3-year-follow-ups. Psychosocial development was evaluated using the emotion regulation ability (measured by the Difficulties in Emotion Regulation Scale (61), DERS) across five approximately annual follow-up waves, and day-to-day executive functioning summary score (measured by Barkley Deficits in Executive Functioning Scale (62), BDEFS) at the 3-year-and 6-year-follow-ups.

#### Concurrent and follow-up assessments for transdiagnostic measures of psychiatric diagnoses

Participants’ current and follow-up psychiatric diagnoses were assessed using a parent-reported and computerized version of the Kiddie Schedule for Affective Disorders and Schizophrenia for DSM-5 (KSADS-5). We calculated the cumulative numbers of KSADS-5 diagnoses at baseline and 2-year-follow-up to derive both the transdiagnostic status and transitions. For analyses regarding the status transition of transdiagnosis, participants were categorized into four subgroups based on whether they met criteria for psychiatric disorders at two time points. These subgroups are: Healthy-persistent (no psychiatric diagnosis for both visits; *N*_Healthy–persistent_ = 3,793), Disorder-new-onset (defined as no diagnosis at baseline but one or more diagnoses at 2-year follow-up; *N*_Disorder–new–onset_ = 569), Disorder-remitted (≥1 psychiatric diagnosis at baseline, but no diagnosis at 2-year follow-up; *N*_Disorder–remitted_ = 874), and Disorder-persistent (≥1 psychiatric diagnosis for both visits; NPP = 357).

*Measures recent adverse childhood experiences across two years*. Recent ACEs score was quantified using the increase in Adverse Childhood Experiences (ACE) scores from baseline to 2-year follow-up. ACE scores were computed based on child and parent reports of potentially traumatic life events, following our previous study (63).

### Regularized multi-set canonical correlation (rmCCA) analysis

To examine the inter-relationships between WM developmental levels, environment and behaviors, we conducted a regularized multi-set canonical correlation analysis (rmCCA) using the *rgcca* package in R (4.6.0). rmCCA extends two-way CCA to accommodate more than two sets of features and identifies the canonical variates. Moreover, rmCCA improves orthogonality among canonical variates compared to conventional sCCA (64). Prior to analysis, all features within each data set were transformed using the *bestNormalize* package in R, based on their distributional properties, and subsequently residualized for age, age^2^ and sex effects. Given the substantially larger number of features in the environmental domain relative to the other two sets, a dimension reduction procedure was applied to the environmental feature set. Multi-collinearity among environmental features was addressed by exploratory factor analyses. The number of factors was determined using Horn’s parallel analysis (factoring method: *minres*; 100 iterations), implemented via the *paran* package in R. This approach compares the observed eigenvalues with those obtained from randomly generated datasets of the same size, thereby accounting for sampling error-induced inflation of eigenvalues. Fifty-eight factors with adjusted eigenvalues greater than one were retained for subsequent analyses (see Supplementary Figure S8 and Data S2). To derive factor scores, an oblique rotation (*oblimin*) was applied. Unlike orthogonal rotations, oblique rotation allows correlations among factors, thereby preserving the inherently interdependent structure of the exposome. To further account for the imbalance in dimensionality across the three data domains, we weighted each view by dividing the square root of the number of variables (65).

The whole CCA framework was build based on the previous sparse two-way CCA analysis procedure by Xia et. al (66). First, the three L1 regularization parameters of the WM developmental, exposomic and behavioral sets were optimized through grid search procedure. The regularization parameters ranged from 0.1 to 1 in 0.1 steps, resulting in 1,000 combinations of the testing parameters. After randomly resampling one-thirds of the sample for ten times, the parameter combination yielding the largest average inner variance explained (*AVE_inner_*) for the first canonical variate on the resampled datasets was used for the further rmCCA analysis in the full sample. To test the significance of global canonical covariance of each canonical variate, we applied a permutation testing procedure (1,000 iterations) to create the null distributions of *AVE_inner_* values for the first 10 canonical variates. During each interaction, we randomly shuffled the subject orders of both the behavioral and environmental sets and kept the WM set constant. The *p* value of each canonical variate was computed by the proportion of the permuted *AVE_inner_* values higher than the original *AVE_inner_* value and corrected by FDR. See Supplementary Figure S1A for the schematic workflow for the permutation tests. Next, we used the bootstrapping (5,000 resamples) to test which features consistently contribute to each canonical variate. During each iteration, two-thirds of the sample were randomly selected, and the remained one-third sample were replaced from the selected sample. As bootstrapping can induce arbitrary orientation and reordering of the canonical modes, we matched the canonical modes derived from the resampling to be consistent with the ordering and the orientation of the original canonical modes. The loading of each individual environmental feature is obtained by multiplying its normalized factor loading by the corresponding rmCCA loading. The feature was considered as significantly consistent for each mode if its two-sided 95% confidence interval did not include zero based on the generated bootstrap distribution.

### Statistical Analysis

The above rmCCA analysis identified two distinct latent dimensions capturing covarying patterns of environment and tract-specific development associated with behaviors during preadolescence. We then tested how differential exposome factors and the related WM development tended to contribute to the cognitive and social development up to six years later.

*Associations of baseline variate scores and follow-up cognitive/social performance*. We conducted univariate analyses using linear mixed effects (LME) models to examine associations (Model 1) between each baseline canonical variate (CV) score (exposome and WM data sets) and each follow-up measure. Models included age and sex as fixed-effect covariates and random intercepts for study site and family ID. Analyses were conducted using the *lmerTest* in R. We report the t values for the CV terms, together with FDR corrected P values.

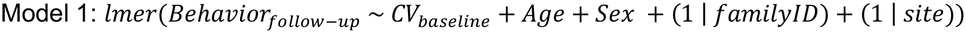

*Associations of baseline variate scores and transdiagnostic status concurrently and at follow-ups*. Participants were categorized into three groups based on the cumulative number of KSADS-5 diagnoses (0, 1, and ≥2 diagnoses) at baseline and at 2-year-follow-up. LME analyses were used to test whether baseline CV scores differed across these groups. We further applied LME models to compare baseline CV scores across longitudinal KSADS-5 diagnostic transition groups (Healthy-persistent, Disorder-new-onset, Disorder-remitted). All models included age and sex as fixed-effect covariates and random intercepts for study site and family ID. Additional models included interactions between sex and diagnostic status or transition group to examine whether these associations differed by sex.

*Moderation analysis*. To test whether the relationship between the changes of both recent ACEs (*rACEs*) and transdiagnostic psychiatric risk (KSADS-5 diagnoses) across two years was moderated by latent environmental or WM developmental profiles, we included two-way interaction terms (*rACEs*: *CV*) in the Model 2. Age, sex, baseline number of K-SADS-5 diagnoses, and baseline ACE score were included as fixed-effect covariates, with random intercepts for study site and family ID.

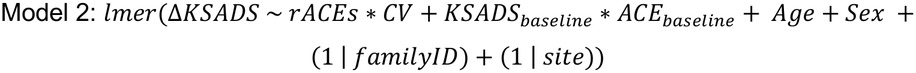

Next, we examined sex-specific moderation effect by testing three-way interaction (*rACEs*: *CV*: *Sex*) to evaluate whether environmental or neurodevelopmental profiles differentially modified these associations by sex.

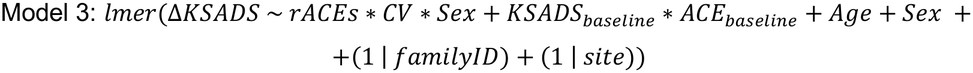

Furthermore, to identify environmental risk-dependent WM makers of resilience, we performed four-way interaction among two environmental variate scores, recent ACEs and the canonical variate WM scores (*CV*_*WM*_), in relation to the cumulative number of KSADS-5 diagnosis at 2-y-followup, in Model 4:

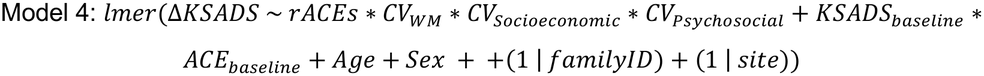

## Data Availability

Participant-level data from the ABCD Study are available through the NIH Brain Development Cohorts Data Use Certification (DUC) process.

https://www.nbdc-datahub.org/

## Acknowledgments

This work was supported by the Intramural Research Program of the National Institute on Drug Abuse, National Institutes of Health (NIH) and utilized the computational resources of the NIH HPC Biowulf cluster (https://hpc.nih.gov). The contributions of the NIH authors were made as part of their official duties as NIH federal employees, are in compliance with agency policy requirements, and are considered Works of the United States Government. However, the findings and conclusions presented in this paper are those of the author(s) and do not necessarily reflect the views of the NIH or the U.S. Department of Health and Human Services.

The authors used data from the Adolescent Brain Cognitive Development Study (ABCD, https://abcdstudy.org/). The ABCD Study, held in the NIH Brain Development Cohorts Data Sharing Platform (https://www.nbdc-datahub.org/), is a multisite, longitudinal study designed to recruit more than 10,000 children aged 9-10 and follow them over 10 years into early adulthood. The ABCD Study is supported by the NIH and additional federal partners under award numbers U01DA041048, U01DA050989, U01DA051016, U01DA041022, U01DA051018, U01DA051037, U01DA050987, U01DA04 1174, U01DA041106, U01DA041117, U01DA041028, U01DA041134, U01DA050988, U01DA051039, U01DA041156, U01DA041025, U01DA 041120, U01DA051038, U01DA041148, U01DA041093, U01DA041089, U24DA041123, and U24DA041147. A full list of supporters is available at https://abcdstudy.org/federal-partners.html. ABCD Consortium investigators designed and implemented the study and/or provided data but did not necessarily participate in the analysis or writing of this report. This manuscript reflects the views of the authors and may not reflect the opinions or views of the NIH or ABCD Consortium investigators.

Additional support for this work was made possible from NIEHS R01-ES032295 and R01-ES031074. This project was also supported by Award Number 2017-MU-CX-0044, awarded by the National Institute of Justice, Office of Justice Programs, U.S. Department of Justice with additional funding from the Centers for Disease Control and Prevention, U.S. Department of Health and Human Services. The opinions, findings, and conclusions or recommendations expressed in this publication/program/exhibition are those of the author(s) and do not necessarily reflect those of the Department of Justice or the Department of Health and Human Services.

## Author Contributions

Conceptualization: DW, CJH, BJS, XX, LM, AJ, TJR, YY. Methodology: DW, XX, TJR. Investigation: DW, CJH, XX, LM, AJ, TJR, YY. Visualization: DW. Supervision: YY. Writing—original draft: D.W., Y.Y., C.J.H. Writing—review & editing: DW, CJH, BJS, XX, LM, HG, TZ, NK, HZ, JH, HL, AJ, TJR, YY.

## References

1. C. A. Nelson, E. F. Sullivan, V. Valdes, Early adversity alters brain architecture and increases susceptibility to mental health disorders. Nat Rev Neurosci 10.1038/s41583-025-00948-9 (2025).

2. E. A. Crone, R. E. Dahl, Understanding adolescence as a period of social-affective engagement and goal flexibility. Nat Rev Neurosci 13, 636–650 (2012).

3. D. Rakesh, S. Whittle, M. A. Sheridan, K. A. McLaughlin, Childhood socioeconomic status and the pace of structural neurodevelopment: accelerated, delayed, or simply different? Trends Cogn Sci 27, 833–851 (2023).

4. D. G. Gee, L. M. Sisk, E. M. Cohodes, N. V. Bryce, Leveraging the science of stress to promote resilience and optimize mental health interventions during adolescence. Nat Commun 13, 5693 (2022).

5. Z. A. Bhutta, S. Bhavnani, T. S. Betancourt, M. Tomlinson, V. Patel, Adverse childhood experiences and lifelong health. Nat Med 29, 1639–1648 (2023).

6. N. Vaidya, A. F. Marquand, F. Nees, S. Siehl, G. Schumann, The impact of psychosocial adversity on brain and behaviour: an overview of existing knowledge and directions for future research. Mol Psychiatry 29, 3245–3267 (2024).

7. K. N. Thakkar et al., Moving from risk to resilience in psychosis research. Nature Reviews Psychology 2, 537–555 %@ 2731-0574 (2023).

8. G. C. George, S. A. Heyn, J. D. Russell, T. J. Keding, R. J. Herringa, Parent Psychopathology and Behavioral Effects on Child Brain-Symptom Networks in the ABCD Study. J Am Acad Child Adolesc Psychiatry 63, 1024–1034 (2024).

9. S. Collishaw et al., Resilience to adult psychopathology following childhood maltreatment: evidence from a community sample. Child Abuse Negl 31, 211–229 (2007).

10. C. Sampaio-Baptista, H. Johansen-Berg, White Matter Plasticity in the Adult Brain. Neuron 96, 1239–1251 (2017).

11. J. K. Knowles, A. Batra, H. Xu, M. Monje, Adaptive and maladaptive myelination in health and disease. Nat Rev Neurol 18, 735–746 (2022).

12. O. de Faria, Jr., et al., Periods of synchronized myelin changes shape brain function and plasticity. Nat Neurosci 24, 1508–1521 (2021).

13. E. L. Dennis et al., Altered white matter microstructural organization in posttraumatic stress disorder across 3047 adults: results from the PGC-ENIGMA PTSD consortium. Mol Psychiatry 26, 4315–4330 (2021).

14. E. Roy et al., Differences in educational opportunity predict white matter development. Dev Cogn Neurosci 67, 101386 (2024).

15. L. Whitmore, D. Beck, Current challenges and future directions for brain age prediction in children and adolescents. Nat Commun 16, 7771 (2025).

16. D. Wang et al., Deviation in development of dorsal association tracts during preadolescence links to concurrent and future cognitive performance and transdiagnostic psychopathology. Nat Commun 17 (2026).

17. M. A. Sheridan et al., Early deprivation alters structural brain development from middle childhood to adolescence. Sci Adv 8, eabn4316 (2022).

18. L. Lim, H. Howells, J. Radua, K. Rubia, Aberrant structural connectivity in childhood maltreatment: A meta-analysis. Neurosci Biobehav Rev 116, 406–414 (2020).

19. H. Huang, T. Gundapuneedi, U. Rao, White matter disruptions in adolescents exposed to childhood maltreatment and vulnerability to psychopathology. Neuropsychopharmacology 37, 2693–2701 (2012).

20. J. Bick, C. A. Nelson, Early Adverse Experiences and the Developing Brain. Neuropsychopharmacology 41, 177–196 (2016).

21. H. Robinson et al., The effect of the “exposome” on developmental brain health and cognitive outcomes. Neuropsychopharmacology 51, 169–184 (2026).

22. S. Genon, A. Ibanez, M. Tahmasian, S. B. Eickhoff, Linking the exposome to the brain-behaviour phenotype. Nat Rev Neurosci 10.1038/s41583-026-01049-x (2026).

23. A. Ibanez et al., Neuroecological links of the exposome and One Health. Neuron 112, 1905–1910 (2024).

24. N. D. Volkow et al., The conception of the ABCD study: From substance use to a broad NIH collaboration. Dev Cogn Neurosci 32, 4–7 (2018).

25. C. Cardenas-Iniguez et al., Building towards an adolescent neural urbanome: Expanding environmental measures using linked external data (LED) in the ABCD study. Dev Cogn Neurosci 65, 101338 (2024).

26. N. E. Holz et al., Early Social Adversity, Altered Brain Functional Connectivity, and Mental Health. Biol Psychiatry 93, 430–441 (2023).

27. D. A. Hackman, M. J. Farah, Socioeconomic status and the developing brain. Trends Cogn Sci 13, 65–73 (2009).

28. D. A. Hackman, M. J. Farah, M. J. Meaney, Socioeconomic status and the brain: mechanistic insights from human and animal research. Nat Rev Neurosci 11, 651–659 (2010).

29. D. Wang et al., Structural Fingerprinting of the Frontal Aslant Tract: Predicting Cognitive Control Capacity and Obsessive-Compulsive Symptoms. J Neurosci 43, 7016–7027 (2023).

30. F. C. Yeh et al., Quantifying Differences and Similarities in Whole-Brain White Matter Architecture Using Local Connectome Fingerprints. PLoS Comput Biol 12, e1005203 (2016).

31. J. M. Nagata et al., Sociodemographic Correlates of Contemporary Screen Time Use among 9-and 10-Year-Old Children. J Pediatr 240, 213–220 e212 (2022).

32. B. Zong, L. Li, Y. Cui, W. Shi, Effects of outdoor activity time, screen time, and family socioeconomic status on physical health of preschool children. Front Public Health 12, 1434936 (2024).

33. S. Marek et al., Patterns of brain-wide associations reflect socioeconomics. Science 392, eaee6213 (2026).

34. A. Ursache, K. G. Noble, N. Pediatric Imaging, S. Genetics, Socioeconomic status, white matter, and executive function in children. Brain Behav 6, e00531 (2016).

35. S. Carozza, I. Kletenik, D. Astle, L. Schwamm, A. Dhand, Whole-brain white matter variation across childhood environments. Proc Natl Acad Sci U S A 122, e2409985122 (2025).

36. K. L. Bottenhorn et al., Adolescent Neurodevelopmental Variance Across Social Strata. JAMA Netw Open 7, e2410441 (2024).

37. Y. M. Ulrich-Lai, J. P. Herman, Neural regulation of endocrine and autonomic stress responses. Nat Rev Neurosci 10, 397–409 (2009).

38. D. C. Dean, 3rd et al., Association of Prenatal Maternal Depression and Anxiety Symptoms With Infant White Matter Microstructure. JAMA Pediatr 172, 973–981 (2018).

39. E. L. Goldwaser et al., White matter in prolonged glucocorticoid response to psychological stress in schizophrenia. Neuropsychopharmacology 46, 2312–2319 (2021).

40. H. I. Sheikh et al., Links between white matter microstructure and cortisol reactivity to stress in early childhood: evidence for moderation by parenting. Neuroimage Clin 6, 77–85 (2014).

41. R. Green et al., Predictors of Substance Use Initiation by Early Adolescence. Am J Psychiatry 181, 423–433 (2024).

42. E. D. Hill et al., Prediction of mental health risk in adolescents. Nat Med 31, 1840–1846 (2025).

43. J. Dubois et al., MRI of the Neonatal Brain: A Review of Methodological Challenges and Neuroscientific Advances. J Magn Reson Imaging 53, 1318–1343 (2021).

44. Y. Li et al., Multiplexed magnetic resonance imaging. Nature 10.1038/s41586-026-10475-x (2026).

45. A. S. Anderson et al., Adverse childhood experiences, parenting, and socioeconomic status: Associations with internalizing and externalizing symptoms in adolescence. Child Abuse Negl 125, 105493 (2022).

46. C. T. Udeh-Momoh et al., Resilience and brain health in global populations. Nat Med 31, 2518–2531 (2025).

47. D. Rakesh, P. A. Lee, A. Gaikwad, K. A. McLaughlin, Annual Research Review: Associations of socioeconomic status with cognitive function, language ability, and academic achievement in youth: a systematic review of mechanisms and protective factors. J Child Psychol Psychiatry 66, 417–439 (2025).

48. H. Eschenbeck, C.-W. Kohlmann, A. Lohaus, Gender differences in coping strategies in children and adolescents. Journal of individual differences 28, 18–26 (2007).

49. A. R. Roeckner, K. I. Oliver, L. A. M. Lebois, S. J. H. van Rooij, J. S. Stevens, Neural contributors to trauma resilience: a review of longitudinal neuroimaging studies. Transl Psychiatry 11, 508 (2021).

50. L. Rachmany et al., Neurodevelopmental effects of early life stress: PFC and cognition. Neurosci Biobehav Rev 186, 106663 (2026).

51. M. C. Cortes-Albornoz, D. P. Garcia-Guaqueta, A. Velez-van-Meerbeke, C. Talero-Gutierrez, Maternal Nutrition and Neurodevelopment: A Scoping Review. Nutrients 13 (2021).

52. B. J. Ellis, W. T. Boyce, Biological Sensitivity to Context. Current Directions in Psychological Science 17, 183–187 (2008).

53. M. P. Herzberg et al., Accelerated maturation in functional connectivity following early life stress: Circuit specific or broadly distributed? Dev Cogn Neurosci 48, 100922 (2021).

54. C. A. Nelson, J. Frankeberger, C. D. Chambers, An introduction to the HEALthy Brain and Child Development Study (HBCD) study. Dev Cogn Neurosci 69, 101441 (2024).

55. H. Garavan et al., Recruiting the ABCD sample: Design considerations and procedures. Dev Cogn Neurosci 32, 16–22 (2018).

56. J. P. Fortin et al., Harmonization of multi-site diffusion tensor imaging data. Neuroimage 161, 149–170 (2017).

57. X. Xiao et al., Brain Functional Connectome Defines a Transdiagnostic Dimension Shared by Cognitive Function and Psychopathology in Preadolescents. Biol Psychiatry 95, 1081–1090 (2024).

58. E. Visoki et al., Classification of Suicide Attempt Risk Using Environmental and Lifestyle Factors in 3 Large Youth Cohorts. JAMA Psychiatry 81, 1020–1029 (2024).

59. A. Brieant et al., Characterizing the dimensional structure of early-life adversity in the Adolescent Brain Cognitive Development (ABCD) Study. Dev Cogn Neurosci 61, 101256 (2023).

60. H. R. Smolker et al., The Emotional Word-Emotional Face Stroop task in the ABCD study: Psychometric validation and associations with measures of cognition and psychopathology. Dev Cogn Neurosci 53, 101054 (2022).

61. K. L. Gratz, L. Roemer, Multidimensional assessment of emotion regulation and dysregulation: Development, factor structure, and initial validation of the difficulties in emotion regulation scale. J Psychopathol Behav 26, 41–54 (2004).

62. A. M. O’Brien, L. R. Kivisto, S. Deasley, J. E. Casey, Executive Functioning Rating Scale as a Screening Tool for ADHD: Independent Validation of the BDEFS-CA. J Atten Disord 25, 965–977 (2021).

63. X. Xiao et al., Modification of Brain Connectome on Association Between Adverse Childhood Experiences and Development of Mental Disorders in Preadolescence. JAMA Netw Open 8, e2533136 (2025).

64. M. Z. Jiang et al., Canonical correlation analysis for multi-omics: Application to cross-cohort analysis. PLoS Genet 19, e1010517 (2023).

65. T. A. Lett et al., Framework for Brain-Derived Dimensions of Psychopathology. JAMA Psychiatry 82, 778–789 (2025).

66. C. H. Xia et al., Linked dimensions of psychopathology and connectivity in functional brain networks. Nat Commun 9, 3003 (2018)

