## Supplementary Information for "Patterns of Childhood Exposomes, White Matter Development and Behaviors Link to Future Cognition and Psychopathology in Youth"

**This PDF file includes:**

Figures S1 to S9
Tables S1 to S4

**Other supporting materials for this manuscript include the following:**

Supplementary Data S1 to S3 (spreadsheet)

### Figures and Tables

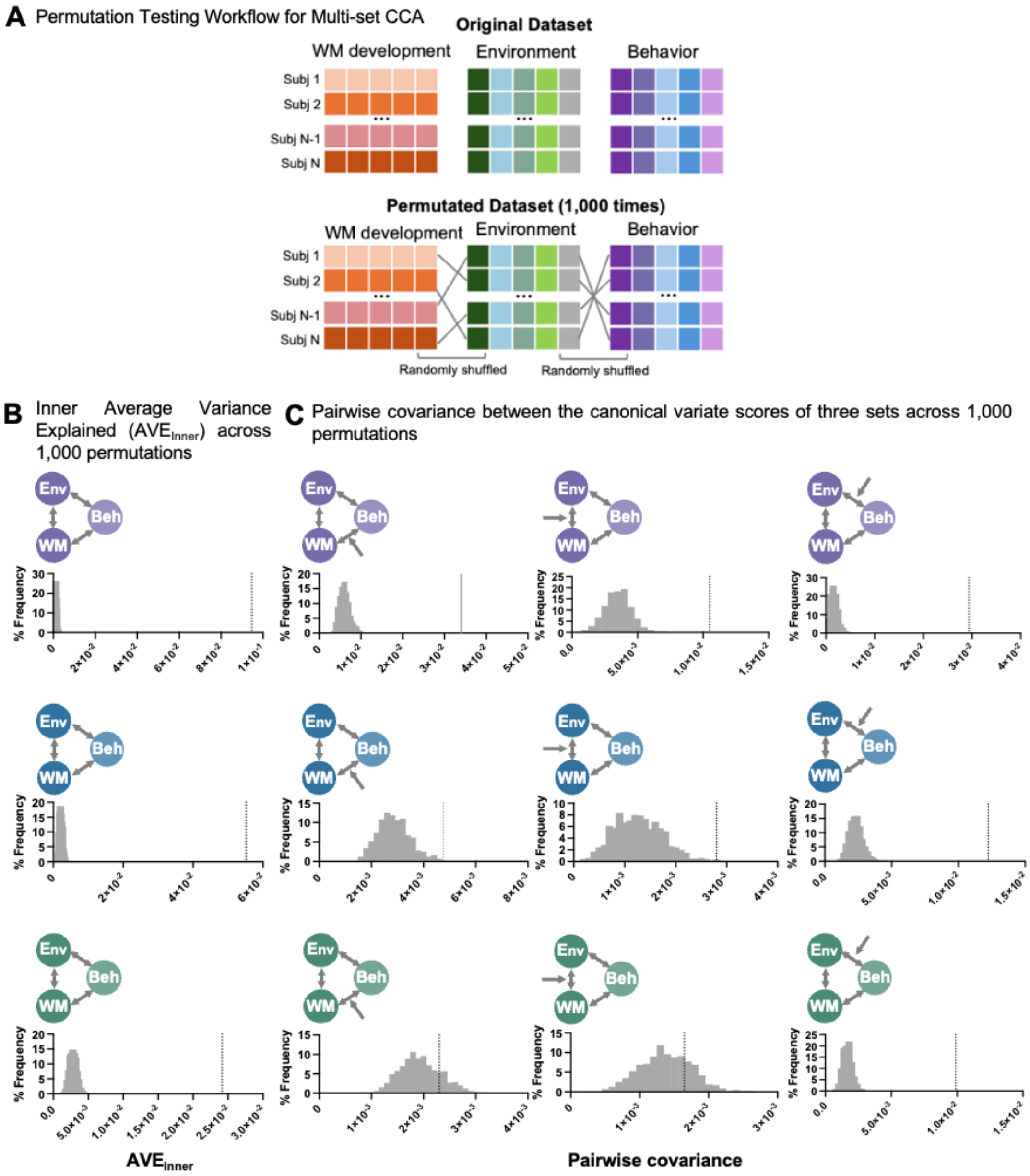


**Supplementary Figure S1. Permutation testing workflow and null distributions for multi-set CCA analysis.**

(A) Permutation framework. Schematic illustration of the permutation procedure (1,000 iterations per test) used to evaluate the statistical significance of multi-set canonical correlations across three domains of white matter (WM), environment (Env) and behavior (Beh). For each permutation, subject labels are randomly shuffled within both Env and Beh sets to disrupt the correspondence of participants' WM features to Env and Beh features.

(B) Global permutation results. Null distributions of inner variance explained ($AVE_{inner}$) for the first three multi-set CCA modes across 1,000 permutations. Gray dotted lines indicate the observed $AVE_{inner}$ values from the original (non-permuted) data.

(C) Permutation results for pairwise covariance between canonical variates. Histograms show the null distributions of pairwise covariance between canonical variate scores for each pair (WM-Env, WM-Beh and Env-Beh) across 1,000 permutations. Network diagrams above each histogram illustrate the corresponding pairs. The vertical gray lines indicate the observed pairwise covariance values from the original data.

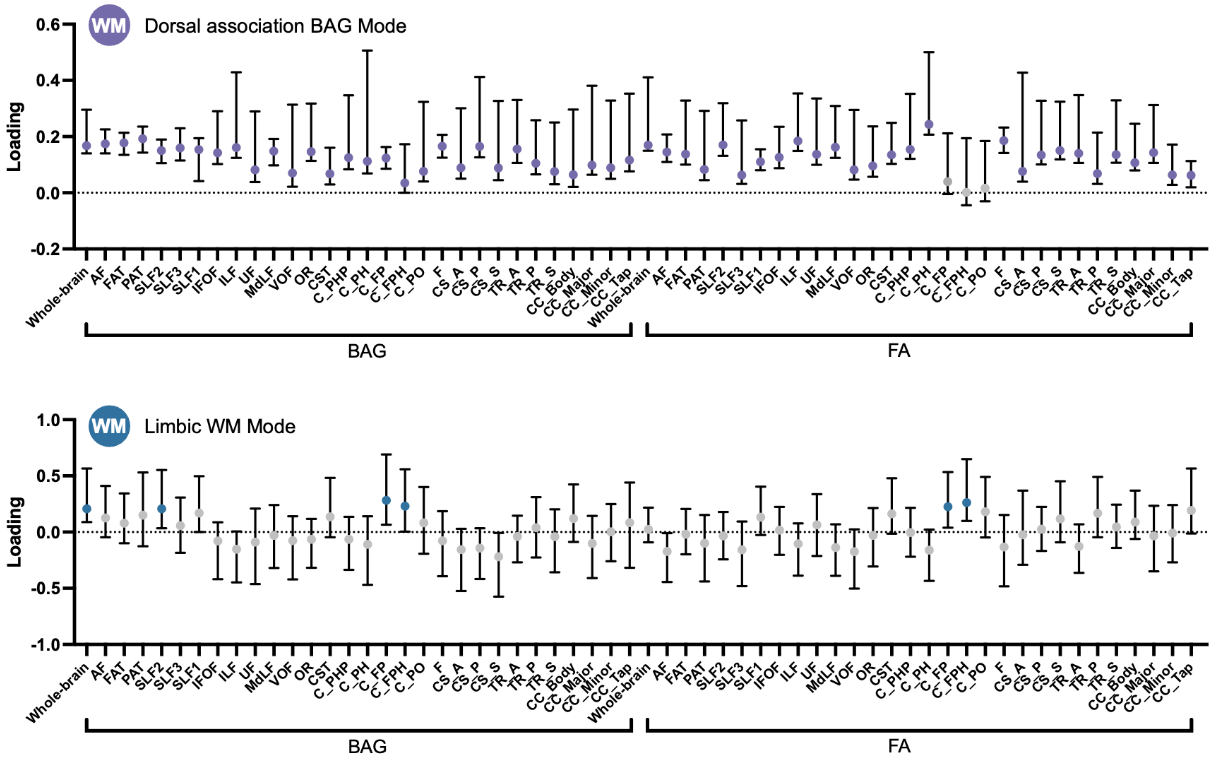


**Supplementary Figure S2. Resampling results to identify stable white matter (WM) features contributing to two linked dimensions.**
Resampling distribution for WM features in each canonical dimension. Each bar represents the 95% confidence interval.

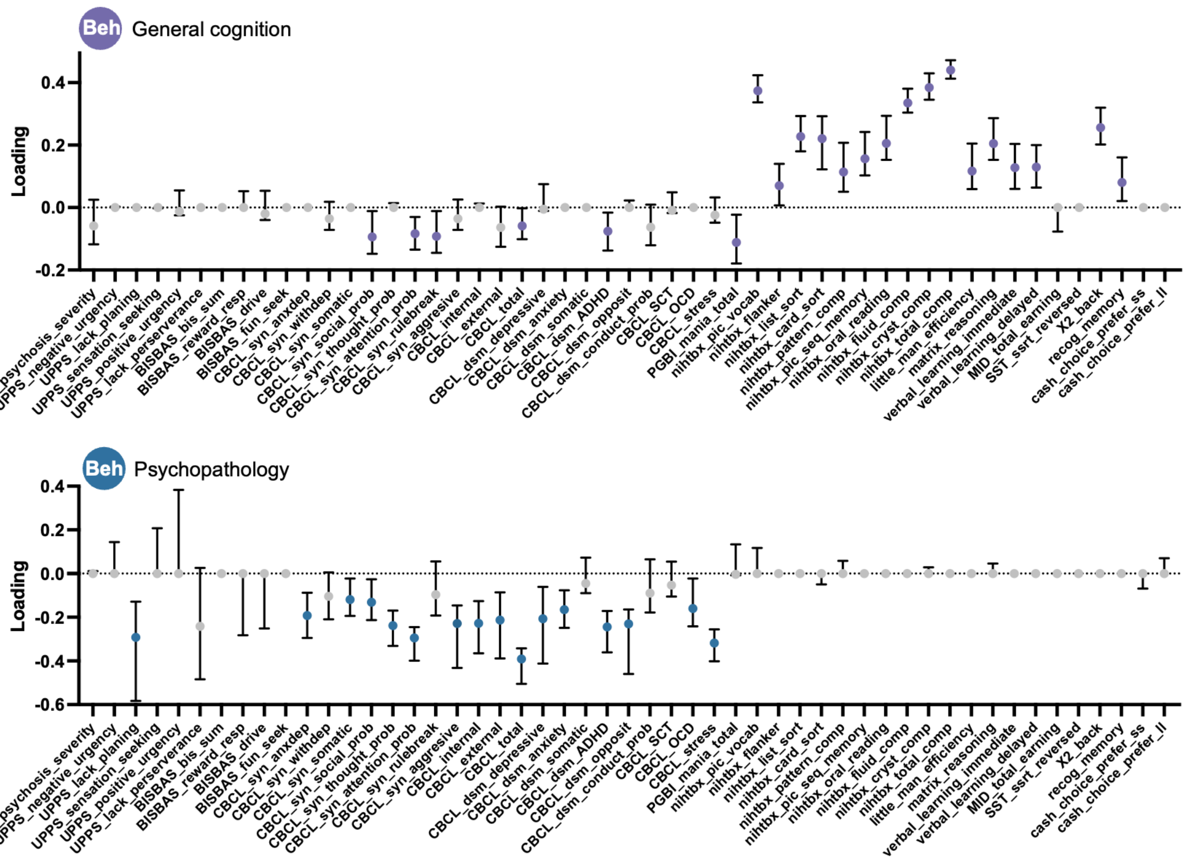


**Supplementary Figure S3. Resampling results to identify stable behavioral (Beh) features contributing to two linked dimensions.**
Resampling distribution for behavioral features in each canonical dimension. Each bar represents the 95% confidence interval.

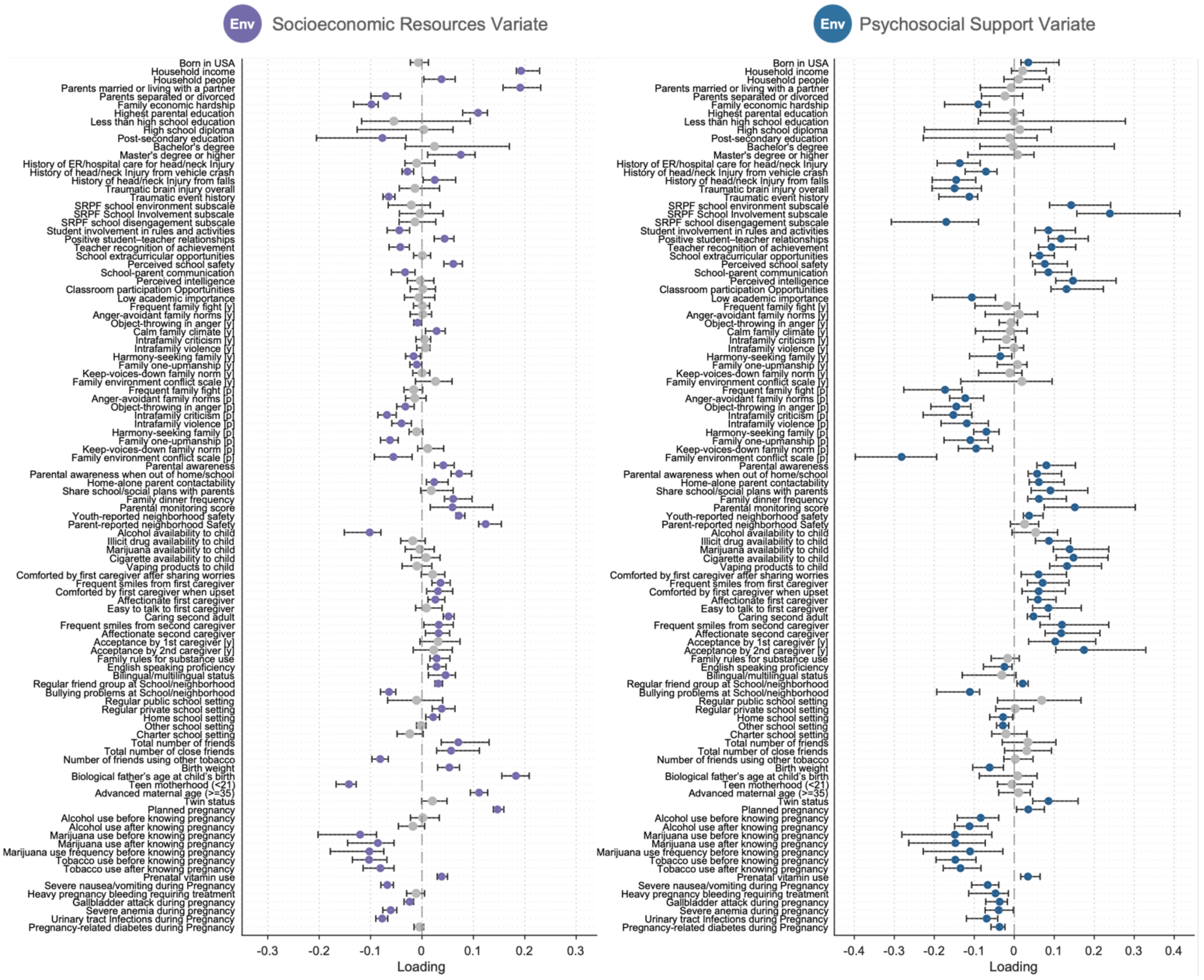


**Supplementary Figure S4. Resampling results to identify stable environmental (Env) features (first 104 features) contributing to two linked dimensions.**

Resampling distribution for environmental features in each canonical dimension. Each bar represents the 95% confidence interval.

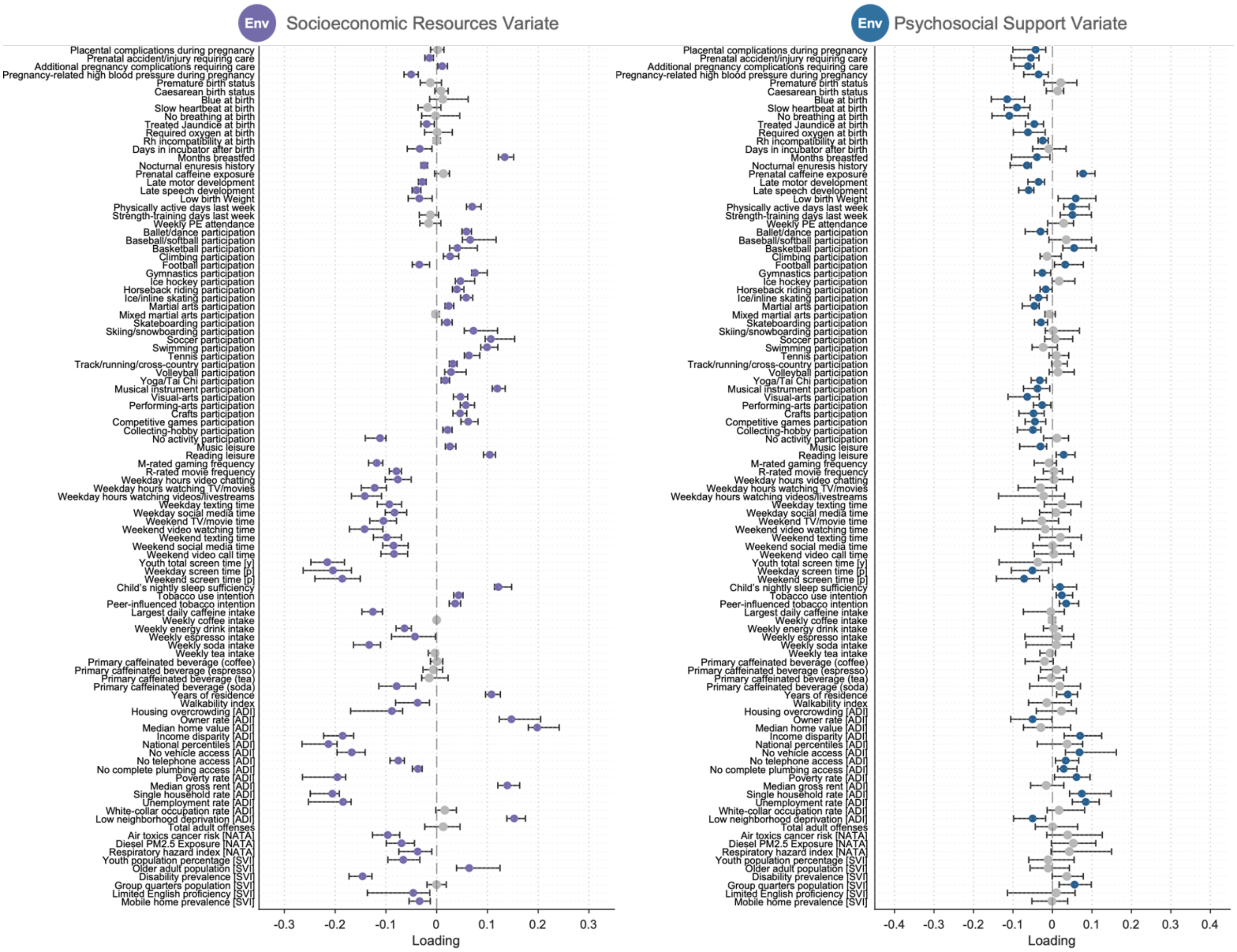


**Supplementary Figure S5. Resampling results to identify stable environmental (Env) features (subsequently 104 features) contributing to two linked dimensions.**
Resampling distribution for environmental features in each canonical dimension. Each bar represents the 95% confidence interval.

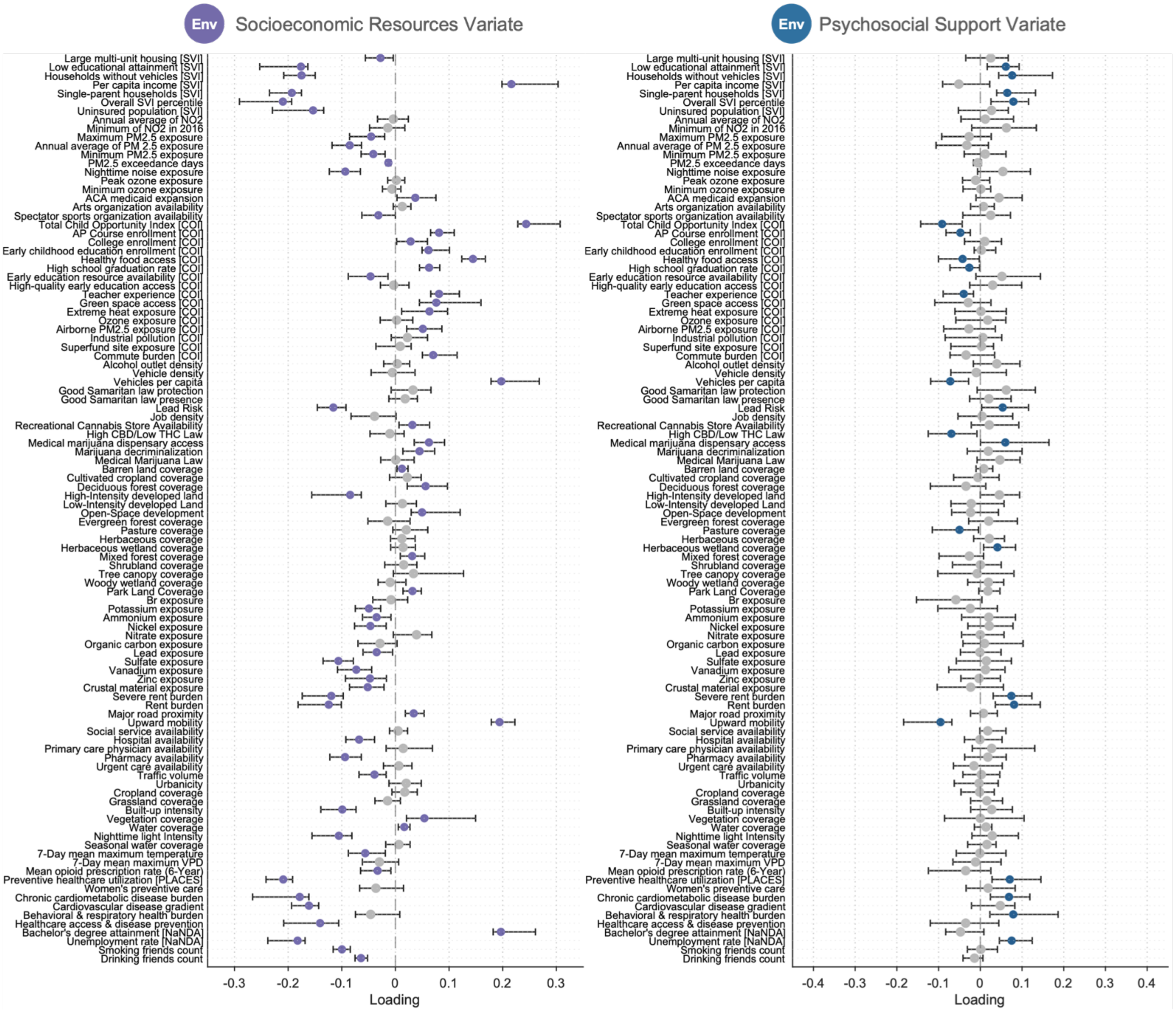


**Supplementary Figure S6. Resampling results to identify stable environmental (Env) features (subsequently 104 features) contributing to two linked dimensions.** Resampling distribution for environmental features in each canonical dimension. Each bar represents the 95% confidence interval.

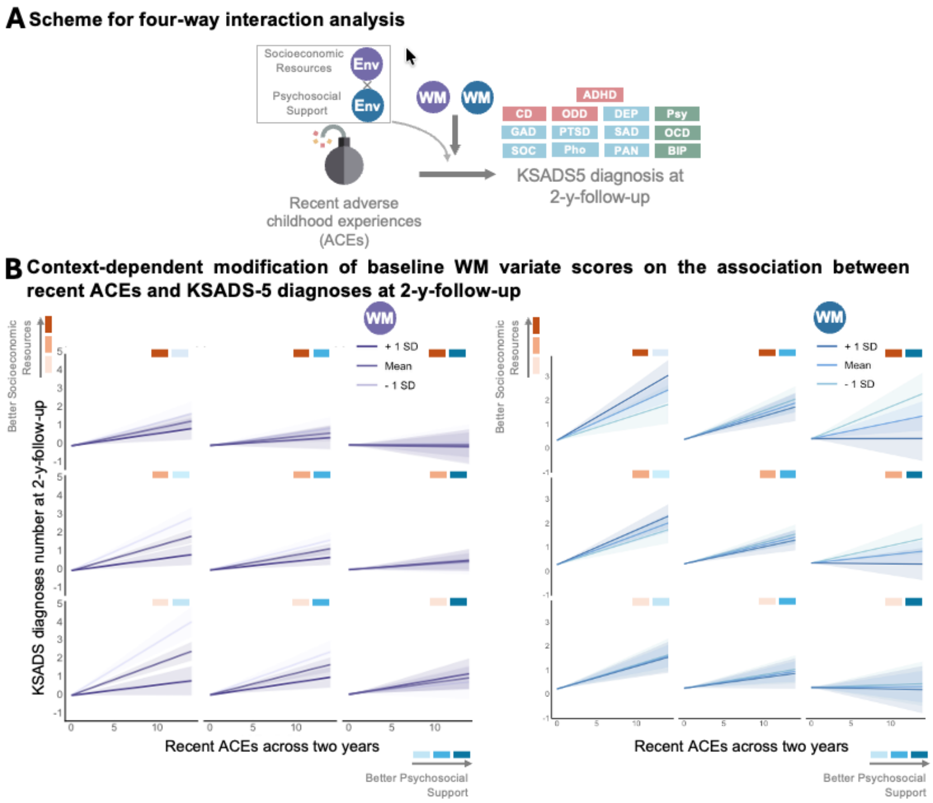


**Supplementary Figure S7**. **Four-way interaction results.**
(A) Schematic illustration of the four-way interaction framework. Environmental dimensions (Socioeconomic Resources and Psychosocial Support Variates) jointly interact with white matter (WM) variate scores to modulate the effect of recent adverse childhood experiences (ACEs) on psychiatric diagnoses.
(B) Context-dependent moderation of WM variate scores under different environmental conditions. The analyses demonstrate that the moderating effect of WM development varies across combinations of environmental exposures. Lines represent simple slopes at +1 SD, mean, and -1 SD of the moderator scores; shaded regions indicate 95% confidence intervals. Abbreviations: KSADS-5, Kiddie Schedule for Affective Disorders and Schizophrenia for DSM-5; rmCCA, regularized multiset canonical correlation analysis.

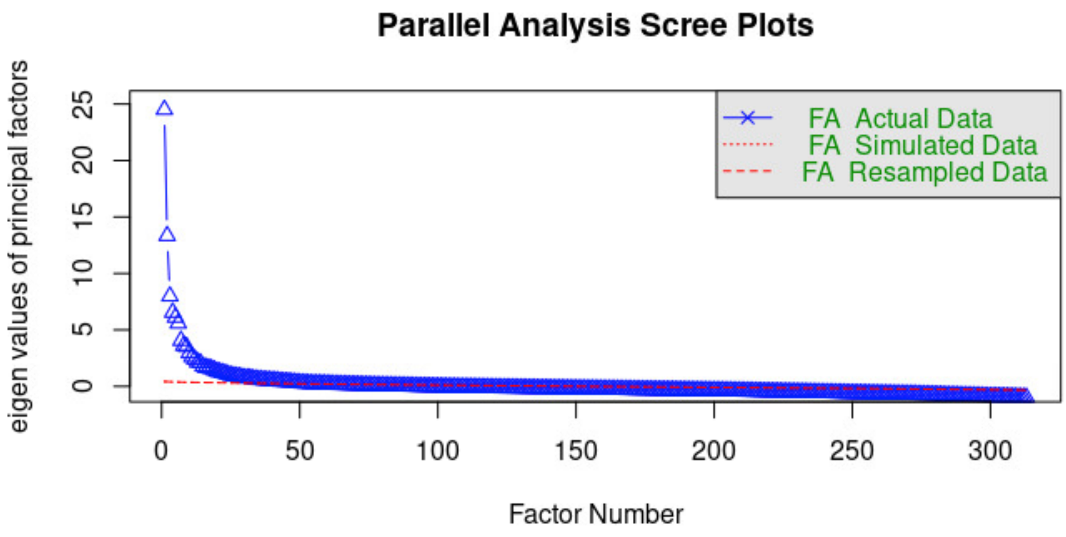


**Supplementary Figure S8. Determination of the optimal number of environmental factors using Horn's parallel analysis.**

Scree plot showing eigenvalues derived from the observed data (blue) and resampled data (red) across factor numbers. The optimal number of factors was defined as the point at which observed eigenvalues exceed those obtained from random data, thereby correcting for sampling error-induced inflation. Based on this criterion, 58 factors were retained for subsequent analyses.

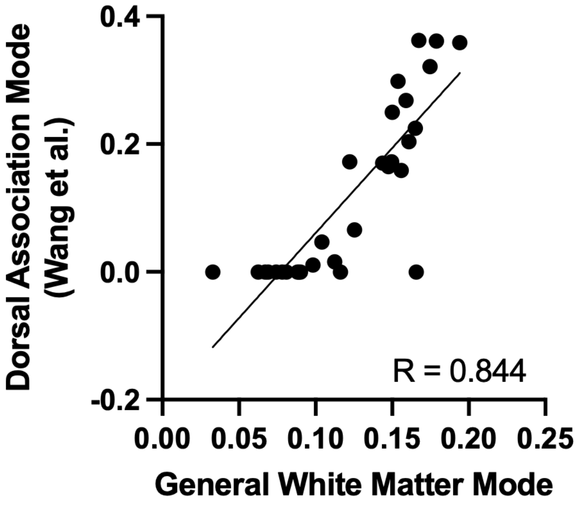


Supplementary Figure S9. **Relationship between tract-specific brain age gap (BAG) loadings in our previous study and the current General WM Variate.** Scatter plot comparing BAG loadings from canonical correlation analysis (CCA) in our previous study with the corresponding BAG loadings for the General white matter (WM) Variate in the current study. Each point represents a WM tract included in both analyses.

**Tables**

**Supplemental Table S1**. Full names and abbreviations of included white matter (WM) tracts.

| Tract Name | Abbreviation |
|---|---|
| Arcuate Fasciculus | AF |
| Frontal Aslant Tract | FAT |
| Parietal Aslant Tract | PAT |
| Superior Longitudinal Fasciculus 1 | SLF1 |
| Superior Longitudinal Fasciculus 2 | SLF2 |
| Superior Longitudinal Fasciculus 3 | SLF3 |
| Inferior Fronto Occipital Fasciculus | IFOF |
| Inferior Longitudinal Fasciculus | ILF |
| Uncinate Fasciculus | UF |
| Middle Longitudinal Fasciculus | MdLF |
| Cingulum Frontal Parahippocampal | C_FPH |
| Cingulum Frontal Parietal | C_FP |
| Cingulum Parahippocampal | C_PH |
| Cingulum Parahippocampal Parietal | C_PHP |
| Cingulum Parolfactory | C_PO |
| Fornix | F |
| Corticospinal Tract | CST |
| Optic Radiation | OR |
| Vertical Occipital Fasciculus | VOF |
| Thalamic Radiation Anterior | TR_A |
| Thalamic Radiation Posterior | TR_P |
| Thalamic Radiation Superior | TR_S |
| Corticostriatal Tract Anterior | CS_A |
| Corticostriatal Tract Posterior | CS_P |
| Corticostriatal Tract Superior | CS_S |
| Corpus Callosum Body | CC_Body |
| Corpus Callosum Forceps Major | CC_Major |
| Corpus Callosum Forceps Minor | CC_Minor |
| Corpus Callosum Tapetum | CC_Tap |

**Supplemental Table S2**. Statistics for the relationship between baseline canonical variate (CV) scores and follow-up cognitive performance

| independent_variable | t_value | coeff | lower_bound | upper_bound | $p_{FDR}$ |
|---|---|---|---|---|---|
| Outcome variable: SMARTE (3-year-follow-up) | | | | | |
| General WM Variate | 9.094 | 0.118 | 0.093 | 0.143 | 6.72E-19 |
| Limbic WM Variate | -3.722 | -0.048 | -0.072 | -0.023 | 0.0004 |
| Socioeconomic Resources Variate | 23.705 | 0.333 | 0.305 | 0.360 | 7.62E-113 |
| Psychosocial Support Variate | -0.554 | -0.008 | -0.035 | 0.019 | 0.650 |
| Outcome variable: SMARTE (5-year-follow-up) | | | | | |
| General WM Variate | 9.611 | 0.120 | 0.095 | 0.144 | 6.53E-21 |
| Limbic WM Variate | -5.063 | -0.062 | -0.086 | -0.038 | 1.40E-06 |
| Socioeconomic Resources Variate | 27.895 | 0.372 | 0.346 | 0.398 | 7.39E-151 |
| Psychosocial Support Variate | -1.110 | -0.015 | -0.040 | 0.011 | 0.332 |
| Outcome variable: School Grade (2-year-follow-up) | | | | | |
| General WM Variate | -6.480 | -0.074 | -0.097 | -0.052 | 3.65E-10 |
| Limbic WM Variate | 2.163 | 0.025 | 0.002 | 0.047 | 0.051 |
| Socioeconomic Resources Variate | -21.209 | -0.265 | -0.289 | -0.240 | 3.09E-94 |
| Psychosocial Support Variate | -7.136 | -0.085 | -0.108 | -0.062 | 4.57E-12 |
| Outcome variable: School Grade (3-year-follow-up) | | | | | |
| General WM Variate | -6.209 | -0.073 | -0.096 | -0.050 | 1.97E-09 |
| Limbic WM Variate | 2.207 | 0.026 | 0.002 | 0.048 | 0.050 |
| Socioeconomic Resources Variate | -21.916 | -0.280 | -0.305 | -0.255 | 2.91E-99 |
| Psychosocial Support Variate | -6.649 | -0.081 | -0.105 | -0.057 | 1.27E-10 |
| Outcome variable: Emotional Stroop task Performance (1-y-follow-up) | | | | | |
| General WM Variate | 7.323 | 0.083 | 0.061 | 0.105 | 1.26E-12 |
| Limbic WM Variate | 0.204 | 0.002 | -0.020 | 0.024 | 0.839 |
| Socioeconomic Resources Variate | 14.405 | 0.176 | 0.152 | 0.200 | 2.39E-44 |
| Psychosocial Support Variate | 1.594 | 0.019 | -0.004 | 0.042 | 0.151 |
| Outcome variable: Emotional Stroop task Performance (3-y-follow-up) | | | | | |
| General WM Variate | 4.212 | 0.055 | 0.030 | 0.081 | 5.76E-05 |
| Limbic WM Variate | -1.198 | -0.016 | -0.041 | 0.010 | 0.294 |
| Socioeconomic Resources Variate | 11.736 | 0.161 | 0.134 | 0.188 | 8.96E-29 |
| Psychosocial Support Variate | 0.295 | 0.004 | -0.023 | 0.031 | 0.796 |
| Outcome variable: Emotional Stroop task Performance (5-y-follow-up) | | | | | |
| General WM Variate | 4.331 | 0.056 | 0.031 | 0.081 | 3.52E-05 |
| Limbic WM Variate | -2.333 | -0.030 | -0.055 | -0.005 | 0.036 |
| Socioeconomic Resources Variate | 9.176 | 0.131 | 0.103 | 0.158 | 5.88E-19 |
| Psychosocial Support Variate | -0.376 | -0.005 | -0.031 | 0.021 | 0.748 |

**Supplemental Table S3**. Statistics for the relationship between baseline canonical variate (CV) scores and follow-up socioemotional measures

| independent_variable | t_value | coeff | lower_bound | upper_bound | $p_{FDR}$ |
|---|---|---|---|---|---|
| Outcome variable: Difficulties in Emotion Regulation (DER; wave 1) | | | | | |
| General WM Variate | -1.579 | -0.037 | -0.083 | 0.009 | 0.153 |
| Limbic WM Variate | -0.708 | -0.016 | -0.062 | 0.029 | 0.559 |
| Socioeconomic Resources Variate | -4.713 | -0.114 | -0.161 | -0.066 | 8.85E-06 |
| Psychosocial Support Variate | -4.500 | -0.105 | -0.150 | -0.059 | 1.77E-05 |
| Outcome variable: Difficulties in Emotion Regulation (DER; wave 2) | | | | | |
| General WM Variate | -0.952 | -0.024 | -0.073 | 0.025 | 0.415 |
| Limbic WM Variate | -1.742 | -0.043 | -0.091 | 0.005 | 0.120 |
| Socioeconomic Resources Variate | -1.633 | -0.043 | -0.095 | 0.009 | 0.144 |
| Psychosocial Support Variate | -5.022 | -0.125 | -0.174 | -0.076 | 1.77E-06 |
| Outcome variable: Difficulties in Emotion Regulation (DER; wave 3) | | | | | |
| General WM Variate | -0.606 | -0.016 | -0.067 | 0.035 | 0.623 |
| Limbic WM Variate | -1.974 | -0.051 | -0.102 | 0.000 | 0.076 |
| Socioeconomic Resources Variate | -0.218 | -0.006 | -0.060 | 0.048 | 0.839 |
| Psychosocial Support Variate | -4.930 | -0.128 | -0.179 | -0.077 | 2.57E-06 |
| Outcome variable: Difficulties in Emotion Regulation (DER; wave 4) | | | | | |
| General WM Variate | -0.375 | -0.010 | -0.062 | 0.042 | 0.748 |
| Limbic WM Variate | -3.963 | -0.105 | -0.156 | -0.053 | 0.0002 |
| Socioeconomic Resources Variate | -0.524 | -0.015 | -0.070 | 0.040 | 0.660 |
| Psychosocial Support Variate | -3.738 | -0.100 | -0.152 | -0.047 | 0.0004 |
| Outcome variable: Difficulties in Emotion Regulation (DER; wave 5) | | | | | |
| General WM Variate | 0.781 | 0.024 | -0.036 | 0.085 | 0.518 |
| Limbic WM Variate | -2.143 | -0.065 | -0.125 | -0.006 | 0.052 |
| Socioeconomic Resources Variate | -1.646 | -0.051 | -0.112 | 0.010 | 0.144 |
| Psychosocial Support Variate | -4.513 | -0.138 | -0.198 | -0.078 | 1.77E-05 |
| Outcome variable: Barkley Deficits in Executive Functioning Scale (BDEFS; 3-y-follow-up) | | | | | |
| General WM Variate | -2.622 | -0.031 | -0.054 | -0.008 | 0.016 |
| Limbic WM Variate | -3.564 | -0.042 | -0.065 | -0.019 | 0.00071 |
| Socioeconomic Resources Variate | -4.920 | -0.065 | -0.091 | -0.039 | 2.57E-06 |
| Psychosocial Support Variate | -20.777 | -0.248 | -0.271 | -0.225 | 7.77E-92 |
| Outcome variable: Barkley Deficits in Executive Functioning Scale (BDEFS; 6-y-follow-up) | | | | | |
| General WM Variate | -1.849 | -0.032 | -0.066 | 0.002 | 0.098 |
| Limbic WM Variate | -2.301 | -0.039 | -0.073 | -0.006 | 0.038 |
| Socioeconomic Resources Variate | -1.504 | -0.029 | -0.066 | 0.009 | 0.173 |
| Psychosocial Support Variate | -13.984 | -0.240 | -0.274 | -0.206 | 3.36E-42 |

**Supplemental Table S4**. Sex difference in the modification of baseline canonical variate (CV) scores (from Limbic white matter [WM], Socioeconomic Resources and Psychosocial Support Modes) on the association between KSADS-5 Diagnoses at year 2, controlling for age, sex, baseline adverse childhood experience and KSADS-5 Diagnoses at baseline. Study site and family were included as the random intercepts.

| Canonical Variate | T value (CV : [recent ACEs]) | p value (CV : [recent ACEs:sex]) |
|---|---|---|
| Limbic WM | 0.918 | 0.359 |
| Socioeconomic Resources | -0.409 | 0.683 |
| Psychosocial Support | -0.176 | 0.861 |

Abbreviations: ACEs, adverse childhood experiences
